# Assessing COVID-19 pandemic excess deaths in Brazil: years 2020 and 2021

**DOI:** 10.1101/2022.07.27.22278096

**Authors:** Saditt Rocio Robles Colonia, Lara Morena Cardeal, Rogério Antonio de Oliveira, Luzia Aparecida Trinca

## Abstract

We estimated the impact of the COVID-19 pandemic on mortality in Brazil for 2020 and 2021 years. We used mortality data (2015–2021) from the Health Ministry, the Brazilian government, to fit linear mixed models for forecasting baseline deaths under non-pandemic conditions. An advantage of the linear mixed model is the flexibility to capture year-trend while dealing with the correlations among death counts over time. Following a specified model-building strategy, estimation of all-cause excess deaths at the country level and stratified by sex, age, ethnicity and region of residence, from March 2020 to December 2021. We also considered the estimation of excess deaths due to specific causes. The estimated all-cause excess deaths was 187 842 (95% PI: 164 122; 211 562, P-Score=16.1%) for weeks 10-53, 2020, and 441 048 (95% PI: 411 740; 470 356, P-Score=31.9%) for weeks 1-52, 2021. P-Score values ranged from 1.4% (RS, South) to 38.1% (AM, North) in 2020 and from 21.2% (AL and BA, Northeast) to 66.1% (RO, North) in 2021. Differences among men (18.4%) and women (13.4%) appeared in 2020 only, and the P-Score values were about 30% for both sexes in 2021. Except for youngsters (*<* 20 years old), all adult age groups were badly hit, especially those from 40 to 79 years old. In 2020, the Indigenous+East Asian population had the highest P-Score (26.2%), and the Black population suffered the greatest impact (34.7%) in 2021. The pandemic impact had enormous regional heterogeneity and substantial differences according to socio-demographic factors, mainly during the first wave, showing that some population strata benefited from the social distancing measures when they could adhere to them. In the second wave, the burden was very high for all but extremely high for some, highlighting that our society must tackle the health inequalities experienced by groups of different socio-demographic status.

## Introduction

By the end of November 2022, Brazil’s coronavirus disease (COVID-19) death toll was 689 665 (nearly 10% of the world confirmed deaths), putting the country among the most affected, behind the USA only [1]. After confirmation of the first cases, around the end of February 2020, the virus spread rapidly from the largest cities to the most vulnerable communities, reaching the whole country by March 2021 [2]. Several factors contributed to the far-beyond disaster in which the Brazilian federal government is blamed to be complicit [3–5]. The President of the Republic chose to follow consistent scientific denialism and assumed a dismissive attitude towards the pandemic. In the middle of chaotic governance, by mid-April 2020, the Supreme Court delegated states/municipalities’ governments the responsibility to rule the pandemic. Nonetheless, efforts were not saved from the federal sphere to undermine the public health responses to COVID-19, assuming a constant confrontation attitude, even for vaccine negotiation and acquisition, which delayed, provoked doubts and disrupted the vaccination process. The consequence was the most drastic. The misinformation spread, the lack of unity and leadership, added to the overcrowded cities, and the difficulties for low-income people accessing social security and engaging in physical distancing all have contributed to the worse [6] [7–11]. Daily COVID-19 mortality reached figures around 1 000 from May to early September 2020. The second pandemic wave started in November, in which daily mortality soared and reached even high figures until July 2021, with the peak, at the end of March, showing more than 3 500 daily COVID-19 deaths [1, 12]. Allied with the already mentioned factors contributing to the virus spread, including mutation and new variants, there is the huge regional diversity concerning health services access and cultural/socio-demographic factors across the country. Although Brazil allegedly has the largest public health system in the world, funded by government budgets, geographical and social inequalities strongly affect access to services [2, 13, 14].

Besides aggregating deaths by the new disease, the pandemic also affected the mortality pattern of other diseases due to changes in social conditions, individual behaviors and, importantly, lack of assistance due to a rather stressed health system. Deaths due to other infections, i.e. influenza, and external causes such as traffic/outdoor accidents and other injuries, are expected to be reduced, mainly in the first months of the pandemic, as a result of restricted social interactions and mobility. On the other hand, deaths due to other causes might have increased due to interruption of treatments, lack of preventive care and other factors. Another problem is the misdiagnose of the cause of death, which leads to under or over-reporting, especially for COVID-19, due to insufficient test capacity, and burdened health and recording systems. [15, 16].

One approach for capturing the total impact, direct and indirect, is by estimating the excess deaths attributable to the pandemic, the difference between the observed and the predicted expectation or baseline deaths, over the same period, had the pandemic not happened [1, 17–19].

Excess deaths in Brazil have been broadly explored using different data sources, locations, periods, stratification factors and methods [7, 10, 13–15, 19–27]. The long-lasting pandemic and mortality data delay means the theme requires constant updating. In this paper, we estimate all-cause excess death for 2020 and 2021 at several aggregation levels, such as in the whole country, by states and considering socio-demographic factors such as sex, age and race/color. For insights to understand excess and/or deficit we also predicted excess deaths due to other specific group causes.

## Materials and methods

### Data

We obtained the publicly available data (2010-2021) from the Mortality Information System (SIM) [12], Ministry of Health, the Brazilian Government, on September 27, 2022. Each SIM record refers to an unidentified death, including the death date, some socio-demographic factors and the primary cause. The data up to December 2020 are considered consolidated while the data for 2021 are preliminary and are expected to change. Data from 2015 to week nine, 2020 were used for modeling the baseline deaths for the pandemic period, while the data from 2010-2019 were used only for the evaluation of the prediction capabilities of the baseline models. Based on 2016-2019 SIM data, IBGE (Instituto Brasileiro de Geografia e Estatística) estimated under-reporting rates ranging from 0.57% to 5.87% with major North and Northeast regions showing the poorer performances [28]. Other Brazilian mortality data exist, however, SIM is considered to have the best coverage and promotion of awareness and training to the personnel of registry and data processing are improving the quality and coverage of the data over the years.

We grouped deaths by epidemiological week (US CDC definition) at several levels of stratification: country, federation unit, sex, age group, race/color and primary death cause. We coded age in five groups, namely 0-19, 20-39, 40-59, 60-79 and 80 or more years old. The factor race/color has five categories, namely White, Black, Brown, Asian and Indigenous. The last two categories were joined because of their single small share. We used this factor as a surrogate for hardship status (economic, education, opportunities access, including healthcare access) since, for historical reasons, the association between these two factors is very well known [29]. Brazil’s territory is organized into five major regions, each sub-divided into several federation units: 26 states and the Federal District, hereafter states for simplicity. For death cause classification, we used the criteria of [19] and formed eight classes: COVID-19, Other Infectious Diseases, Neoplasms, Cardiovascular Diseases, Respiratory Diseases, Ill-Defined Causes, External Causes and Other Diseases. We note that all COVID-19 deaths recorded in the SIM used the ICD coding B-34.2 (no death has been recorded as U07.1 or U07.2 yet).

For data preparation and handling, we acknowledge enormous benefits from the publicly available program code from [19].

## Methods

We used the linear mixed model (LMM) to predict baseline deaths. In the following we present our reasoning in favor of this method.

The available methods broaden from simple five-year averaging to quite demanding models requiring census data [17, 18, 30–32]. [18] brought to mind, apart from other problems, the data violations on assumptions of some of the approaches. A typical data violation is the implicit correlations of the historical death counts not incorporated in several models. Another point is that some methods fail to account for the specific-year mortality trend, i.e. averaging over years without adjustments. The mixed models are devised for modeling grouped/correlated data [33], capturing correlations within and heterogeneity among groups. The linear case was applied successfully to mortality data for two European countries [18]. The normality assumption is decisive for the simplicity and flexibility of the modeling, allowing the incorporation of a broader class of correlation structures, achieving predictions for specific groups, and attaining explicit expressions for the standard errors of the predictions. The last point is crucial to take into account all uncertainty involved in the forecasts. Furthermore, it does not require information from census population projections. That is not necessarily the case when the modeling includes some non-linear link function and assumes some other non-normal distribution, as in the Generalized Linear Model (GLM) framework. The so-called WHO method for baseline deaths uses a GLM [31], which requires simulations from the fitting to obtain reliable prediction standard errors. Yet, as far as we understand, it does not include correlations.

The criticism of LMM is the underlying normality assumption, but since weekly mortality assumes large values, a good approximation is, generally, attained [7, 18].

Another possible approach is the application of the Generalized Estimation Equations (GEE) formulation [32], which, while considering correlations among the observations, does not require any distributional assumption. The model is devised for grouped/correlated data to estimate the mean profile for the population from which the groups represent a sample. Its drawback, in our opinion, is that it is not devised for predictions. We can use the fitted equation to extrapolate it to future time points and obtain the point predictions. However, the precision of such predictions, considering only the uncertainty on parameter estimates, is not fair because it does not consider the uncertainty related to future observations. Failing accurate estimation of the standard errors of the baseline predictions could result in too narrow prediction intervals and lead to incorrect interpretations of excess deaths. Nonetheless, we used this approach as a base for comparisons to the LMM approach, in terms of point predictions, applied to the Brazilian data.

### Modeling by the linear mixed model

Using historical mortality data, for the LMM framework, each year is considered a cluster or group and weeks within a year are the observational units [18]. Terms of the Fourier Series (FS) captures the cyclic pattern and year random effects capture year trend, such that a basic model is

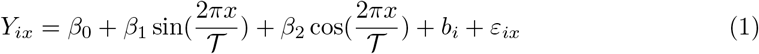

where *Y*_ix_ is the death count in year *i* (*i* = 1, 2, …, *M*) and week *x* (*x* = 1, 2, …, 52), *β*’s are fixed-effect parameters, 𝒯 is the period (of the FS), 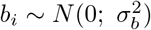 is the year-specific random effect and *ε*_ix_ ∼ *N* (0; *σ*^2^) is the random measurement error. As standard, *b*_i_ and *ε*_ix_ are assumed to be independent. It is possible to include other random effects to account for the heterogeneity of the regression parameters over the years, as can other relevant covariates of fixed effects.

The first documented COVID-19 death in Brazil occurred on March 12, 2020, allowing a non-pandemic period of nine weeks to predict the 2020-specific death trend, relaxing the requirement of census data to capture mortality rates. However, for 2021, the prediction of such a trend is impossible using Eq (1) and we would have to rely on the estimated population mean curve. To obtain more realistic forecasts for 2021 beyond the estimated population mean, which would underestimate the baseline deaths, we have included a linear term of time in the model. Thus, the more general LMM is

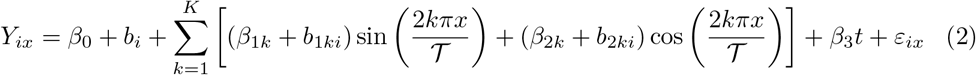

where *K* is the number of terms needed in the FS, *b*_1ki_ and *b*_2ki_ are further random effects to account for year heterogeneity and *t* = 1, 2, …, 269 indicates time points in the series of the data, starting in week 1, 2015 and ending in week nine, 2020. Using this type of model we predicted weekly mortality for 2020 and 2021, aggregated for all-cause deaths, at the country level and stratified by: state, sex, age group and race/color categories. We further obtained baseline values for primary death-cause groups. As the mixed model involves modeling mean and variance-covariance structures, changes in one part might impact the estimates of the other, and care should be taken for fitting a parsimonious model. We outline below the general steps we followed to select a parsimonious model for each stratification.

1. Fit a linear fixed-effects model using Ordinary Least Squares (OLS), considering the years as a blocking factor and the weeks as a qualitative factor. This model, with 58 parameters, is the completest mean model the data allow fitting. Denote it Model 0.
2. Search for some function of *x* (week) that adequately captures the yearly cyclic pattern. In this step, the number of terms (*K*) required in the FS in Eq (2) should be fixed. To estimate the period 𝒯 (or equivalently the frequency 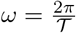), the approach indicated in [33] is used, that is, fit a non-linear model (non-linear least squares) to estimate the *ω* parameter. The non-linear model includes years as a blocking factor, a linear effect of time *t* and *K* fixed, say *K* = 2. Note that such a model captures the year effect but remains partially linear, requiring an initial value for *ω* only. With 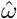 given, fit the linear regression model that incorporates the functional form of *x* and *t* found previously. Note that such a model represents a considerate simplification of Model 0. Test for lack-of-fit of the simpler model. Under the evidence of lack-of-fit, increase *K* and repeat the checking. In most cases, *K* = 1 or 2 resulted in parsimonious fits, but there were cases requiring *K* = 3. The final model in this step is Model 1.
3. Fit the LMM with the mean part found in step 2 and the random effects, one for accounting for years variability and one associated to each element of the FS in Eq (2). Note that once more than one random effect is included in the model, then we have a random vector **b**_i_, with some covariance matrix **D**, such that **b**_i_ ∼ *N* (**0**; **D**) and the structure of **D** must be specified. We started with the most complex structure, i.e., the so-called unstructured pattern, which means any symmetric positive-definite matrix. The fitted model in this step is Model 2.
4. Simplify, if possible, the structure of **D** by declaring **D** diagonal, which means the random effects are uncorrelated. Denote Model 3 the most parsimonious model in this step.
5. Explore other models, possibly dropping random effects based on the magnitude of their variance component estimates and their standard errors, or performing some formal tests. The simpler model, not showing lack-of-fit compared to the more complex one, is kept. Call Model 4 the final model in this step.
6. Update Model 4 by incorporating serial correlation between observations within the same year. That is, the no-serial correlation model assumes *ε*_i_ ∼ *N* (**0**, *σ*^2^**I**). For serial correlation, *V ar*(*ε*_i_) = **R**, where **R** is a non-diagonal symmetric positive-definite matrix. Some usual possibilities are first-order auto-regressive (AR(1)), Gaussian and Spherical correlation patterns [33]. The most parsimonious model is Model 5.
7. Check for further simplification of the random part as some random effects in **b**_i_ might not be relevant after the serial correlation account. Keep the most parsimonious model (Model 6).
8. Check for simplification of the fixed effects (mean model) by applying a backward type selection.
9. Perform a detailed diagnostic analysis of the fit. In case of evidence of violations, some fix may be possible by following the recommendations in [34].

Applications of these steps allowed the selection of a model for baseline death predictions for each stratification we explored. For most cases, one term in the FS was enough to explain the seasonal mortality variation, but there were cases requiring two or three. In particular, for External Causes of death, the FS function did not fit the data. We used a third-order polynomial instead. Often, the serial correlation structure was well-modeled by AR(1).

The estimated equation applied to weeks *x* = 10, 11, …, 53, time points *t* = 270, 271, … 313 and the predicted 2020-specific random effects provided the baseline death forecasts for the pandemic period of the year 2020. Similarly, for the year 2021 forecasts, weeks *x* = 1, 2, … 52 and time points *t* = 314, 315, …, 365 were applied, the difference being that the best predictions of 2021-specific random effects are null. For calculations of standard errors and prediction intervals, we used standard LMM theory (for details, see S1 File).

### Modeling by GEE

For the GEE formulation, the primary death cause stratum (seven in our case) defines the grouping. The model has the three components described below.

1. The link function (the natural choice is the log link since the response is death counts) related to the linear predictor

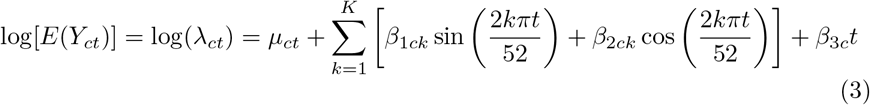

where *λ*_ct_ and *µ*_ct_ are, respectively, the expected death count and the intercept, in stratum *c* (*c* = 1, 2, …, 7) and time point *t*. The other terms follow definitions already stated around Eq (2), however, specific for stratum *c*.
2. The variance function: *V ar*(*Y*_ct_) = *ϕλ*_ct_ whose form declares the variance follows the behavior of the over-dispersed Poisson model.
3. The correlation function specifies the correlation pattern among death counts at distinct time points. Here, we assumed the AR(1) structure.

We note that this is not the usual specification of the GEE approach used to model population mean profiles using a sample of groups or clusters because the linear predictor includes the specific effects for the grouping factor, and the fitted model estimates the mean profile group-specific. In this context, it should be so because there is no meaning in averaging the deaths (or log deaths) among the distinct death causes. By substituting *t* = 270, 271, …, 365 in the fitted linear predictor of Eq (3) and applying exponentiation, a baseline mortality forecast was obtained for each time point and grouping. Aggregation across groups, following the methods in [32], resulted in baseline forecasts for all-cause deaths.

### Prediction Accuracy

To assess the prediction accuracy of both modeling approaches, LMM and GEE, we used the usual measures related to prediction error (for details see S1 File). For this task, we used data from 2010-2019 such that for each year *i* in turn, from 2015 to 2019 (*i* = 1, 2, …, 5), we fitted each model being compared using data from the previous five-year history and the nine first weeks of year *i*. Then, for a year *i*, we obtained forecasts for weeks 10, 11, …, 52 and calculated the prediction error as the difference between the actually observed and the predicted death counts. We conducted this investigation only for the all-cause deaths at the country level.

### Excess deaths

The excess death estimate for each time point is the observed death count minus the predicted baseline. For year summaries and fairer comparisons between strata, we used the P-Score (per capita of excess death in percentage) [35] and the Ratio_EC_ (ratio excess by COVID-19 deaths) [19]. The year-accumulated P-Score is defined as 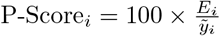 where *E*_i_ is the accumulated excess and 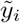 is the accumulated baseline forecast deaths for year *i*. The year-accumulated Ratio_ECi_ is 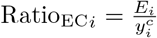 where 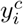 is the annual COVID-19 confirmed deaths. The Ratio_ECi_ measures the COVID-19 pandemic effect on deaths attributable to other causes. Values below one mean that COVID-19 surpassed excess deaths such that there was a deficit in reporting deaths due to other causes, while values above one mean extra deaths due to other causes beyond COVID-19 amounted.

### Computational resources

We used R [36] for all computations (packages: nlme [37] for the LMM, varTestnlme [38] for adjusting p-values when necessary, geepack [39] for GEE model, ggplot2 [40] for the plots). For the fitting diagnosis, we used the R function lmmdiagnostic freely available for download at http://www.ime.usp.br/~jmsinger/lmmdiagnostics.zip.

## Results

### LMM versus GEE

Firstly we show the results comparing the performances of the best fittings we found following the two modeling alternatives presented in the methodology. We concentrate on the forecasting of all-cause baseline deaths at the country level. Fig 1 shows the death count series from the first week of 2015 to week nine of 2020 and the fitted curves by both models. Both models underestimate the yearly peak, with LMM showing a somewhat better performance. LMM also captures the lower spikes that appear consistently at the end/beginning of each year.

**Fig 1.**
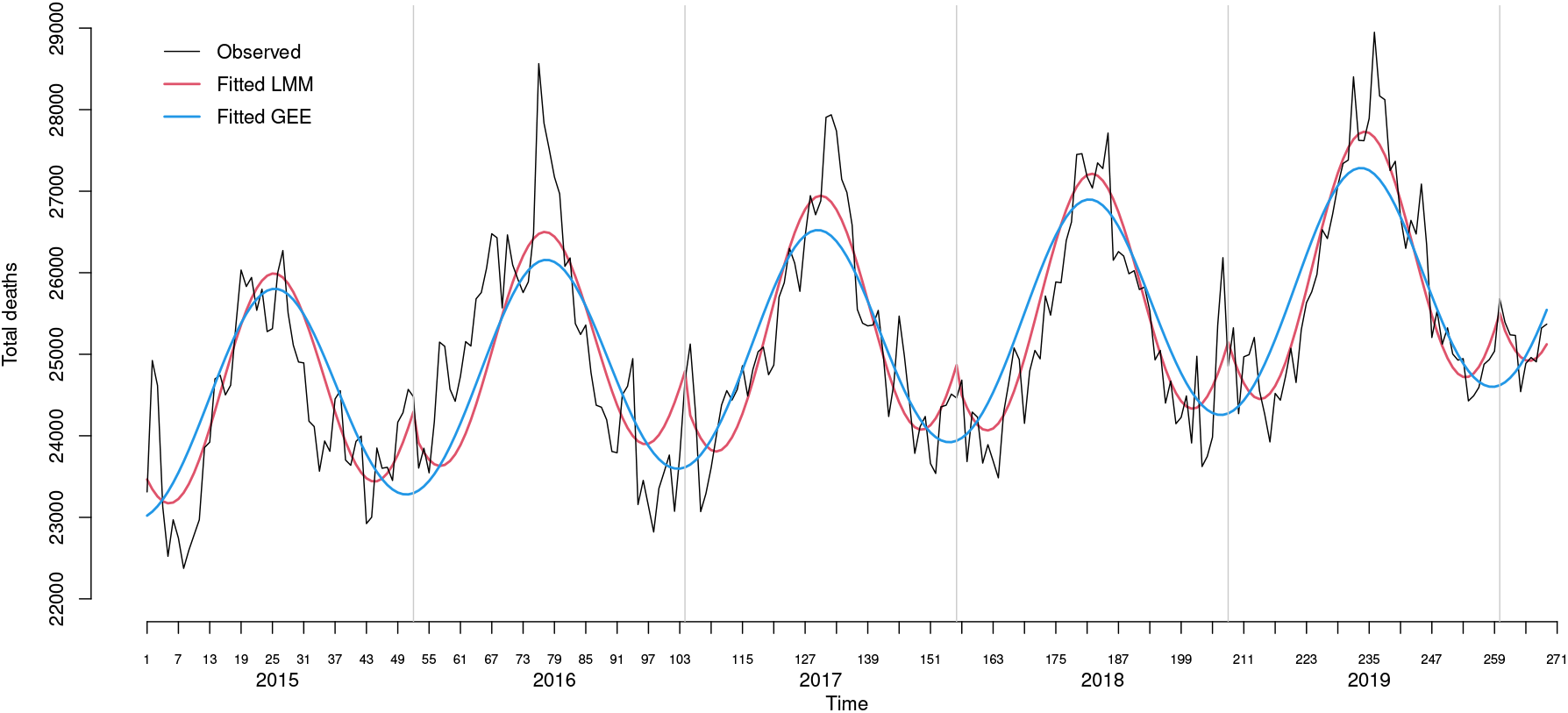
Weekly all-cause mortality in Brazil from week 1, 2015 to week nine, 2020. Recorded and fitted mortality by the LMM and the GEE approach.

Table 1 presents summaries of prediction accuracy measures when using the fitted models to predict the number of deaths for weeks from 10 to 52 for years from 2015 to 2019. The data used to fit the models were always from the previous five-year period plus the first nine weeks of the current year. For comparison, we also included the five-year average performance. LMM and GEE present very similar performances that are, unsurprisingly, much superior to the five-year average method.

**Table 1.** Summaries for accuracy measures under LMM and GEE for forecasting death numbers in future weeks of the year, using data from the previous five-year period up to the ninth week of the current year.

| Model | Summary | Accuracy measures<sup>a</sup> |  |  |  |  |  |
| --- | --- | --- | --- | --- | --- | --- | --- |
|  |  | ME | MAE | RMSE | RRMSE | MAPE | MPE |
| LMM | *min* | -371.52 | 389.69 | 544.88 | 0.32 | 1.45 | -1.51 |
|  | *mean* | 59.92 | 614.14 | 748.92 | 0.45 | 2.42 | 0.15 |
|  | *median* | 189.21 | 521.45 | 622.90 | 0.37 | 2.07 | 0.65 |
|  | *max* | 384.49 | 961.88 | 1145.50 | 0.69 | 3.77 | 1.50 |
| GEE | *min* | -447.29 | 515.05 | 653.42 | 0.38 | 1.95 | -1.83 |
|  | *mean* | -27.31 | 664.32 | 794.63 | 0.48 | 2.62 | -0.21 |
|  | *median* | 59.98 | 646.64 | 763.45 | 0.46 | 2.62 | 0.13 |
|  | *max* | 288.53 | 919.68 | 1091.20 | 0.66 | 3.61 | 1.00 |
| Five-year<br>Average | *min* | 1120.64 | 1131.94 | 1244.81 | 0.74 | 4.39 | 4.35 |
|  | *mean* | 1496.20 | 1498.46 | 1640.46 | 0.99 | 5.86 | 5.85 |
|  | *median* | 1525.41 | 1525.41 | 1593.01 | 0.96 | 5.95 | 5.95 |
|  | *max* | 1865.60 | 1865.60 | 2136.65 | 1.29 | 7.25 | 7.25 |

<sup>a</sup>: ME: mean error; MAE: mean absolute error; RMSE: root mean squared error; RRMSE: relative root mean squared error; MAPE: mean absolute percentage error and MPE: mean percentage error.

The reasonable agreement between the LMM and GEE in point predictions can also be seen in Table 2, although there is a tendency for lower baseline forecasts from the GEE. Given the flexibility of the LMM, allowing explicit formulae for uncertainty estimation around point predictions, we will concentrate, in the following sections, on LMM results.

**Table 2.** Reported, expected and estimated excess/deficit deaths by primary selected cause, accumulated for two periods, weeks 10-53, 2020 and weeks 1-52, 2021.

| Year | Cause | Reported | Expected |  | Excess |  | 95% PI<sup>a</sup> |  | P-Score<sup>a</sup> |
| --- | --- | --- | --- | --- | --- | --- | --- | --- | --- |
|  |  |  | LMM | GEE | LMM | GEE |  |  |  |
| 2020 | All-cause | 1 351 320 | 1 163 478 | 1 136 629 | 187 842 | 189 096 | 164 122 | 211 562 | 16.1 |
| 2020 | COVID-19 | 214 620 |  |  |  |  |  |  |  |
| 2020 | Cardiovascular | 302 999 | 313 349 | 306 593 | -10 350 | -3 594 | -17 757 | -2 943 | -3.3 |
|  | Other Diseases | 269 045 | 271 267 | 265 817 | -2 222 | 3 228 | -9 926 | 5 483 | -0.8 |
|  | Neoplasms | 191 404 | 204 005 | 199 772 | -12 601 | -8 368 | -14 654 | -10 548 | -6.2 |
|  | Respiratory | 125 941 | 140 921 | 137 907 | -14 980 | -11 966 | -21 348 | -8 613 | -10.6 |
|  | External | 122 569 | 124 901 | 119 499 | -2 332 | 3 070 | -6 522 | 1 859 | -1.9 |
|  | Ill-defined | 79 235 | 64 145 | 61 375 | 15 090 | 17 860 | 10 927 | 19 254 | 23.5 |
|  | Other Infect. | 45 507 | 47 304 | 46 365 | -1 797 | -858 | -2 817 | -777 | -3.8 |
| 2021 | All-cause | 1 821 737 | 1 380 689 | 1 382 939 | 441 048 | 438 798 | 411 740 | 470 356 | 31.9 |
| 2021 | COVID-19 | 420 193 |  |  |  |  |  |  |  |
| 2021 | Cardiovascular | 377 828 | 368 705 | 368 135 | 9 123 | 9 693 | -874 | 19 119 | 2.5 |
|  | Other Diseases | 337 057 | 326 048 | 328 318 | 11 009 | 8 739 | 2 435 | 19 582 | 3.4 |
|  | Neoplasms | 233 344 | 246 810 | 248 149 | -13 466 | -14 805 | -15 743 | -11 188 | -5.5 |
|  | Respiratory | 142 975 | 165 266 | 164 740 | -22 291 | -21 765 | -29 439 | -15 144 | -13.5 |
|  | External | 147 003 | 150 870 | 143 440 | -3 867 | 3 563 | -15 713 | 7 979 | -2.6 |
|  | Ill-defined | 100 645 | 80 508 | 73 985 | 20 137 | 26 660 | 11 091 | 29 183 | 25.0 |
|  | Other Infect. | 62 692 | 55 605 | 56 172 | 7 087 | 6 520 | 4 794 | 9 379 | 12.7 |

*a*: based on the LMM model.

### Excess deaths at the country level

Detailed results from our step-by-step strategy to model the baseline deaths from all-cause at the country level is presented in S2 File.

The final fitted baseline model for the year 2020 (*i* = 6) is presented in Eq (4). Two terms of the Fourier series with the estimated frequency 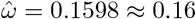, resulting in a period of approximately 39 weeks, were adequate. The best selected structure for **D** was diagonal with square-root variance component estimates given by 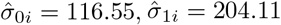 and 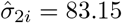. The best structure for **R** was found to be the AR(1), with 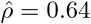 and 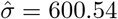. Diagnostics graphs did not reveal any concern about violations of the assumptions of the model (see S1 Fig), reassuring the usefulness of the LMM approach. Thus, for 2020 the predictor equation is

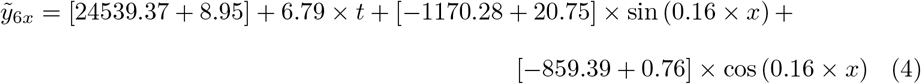

where *x* = 1, 2, …, 53 and *t* = 260, 261, …, 313. The second figure within each set of square brackets in Eq (4) is the prediction of the 2020-specific effect. For 2021, the equation is the same except that year-specific effects are set to zero and *x* runs from 1 to 52 and *t* from 314 to 365. The great impact if we predict for 2020 or 2021 is, therefore, concerning the uncertainty around the predictions (see S1 File) as we will show in the graphs.

Fig 2 shows the observed weekly all-cause mortality for week 1 of 2015 to week 52 of 2021, the baseline forecasts for both years, and the 95% PI for expected mortality plus reported COVID-19 deaths (forecast + COVID-19).

**Fig 2.**
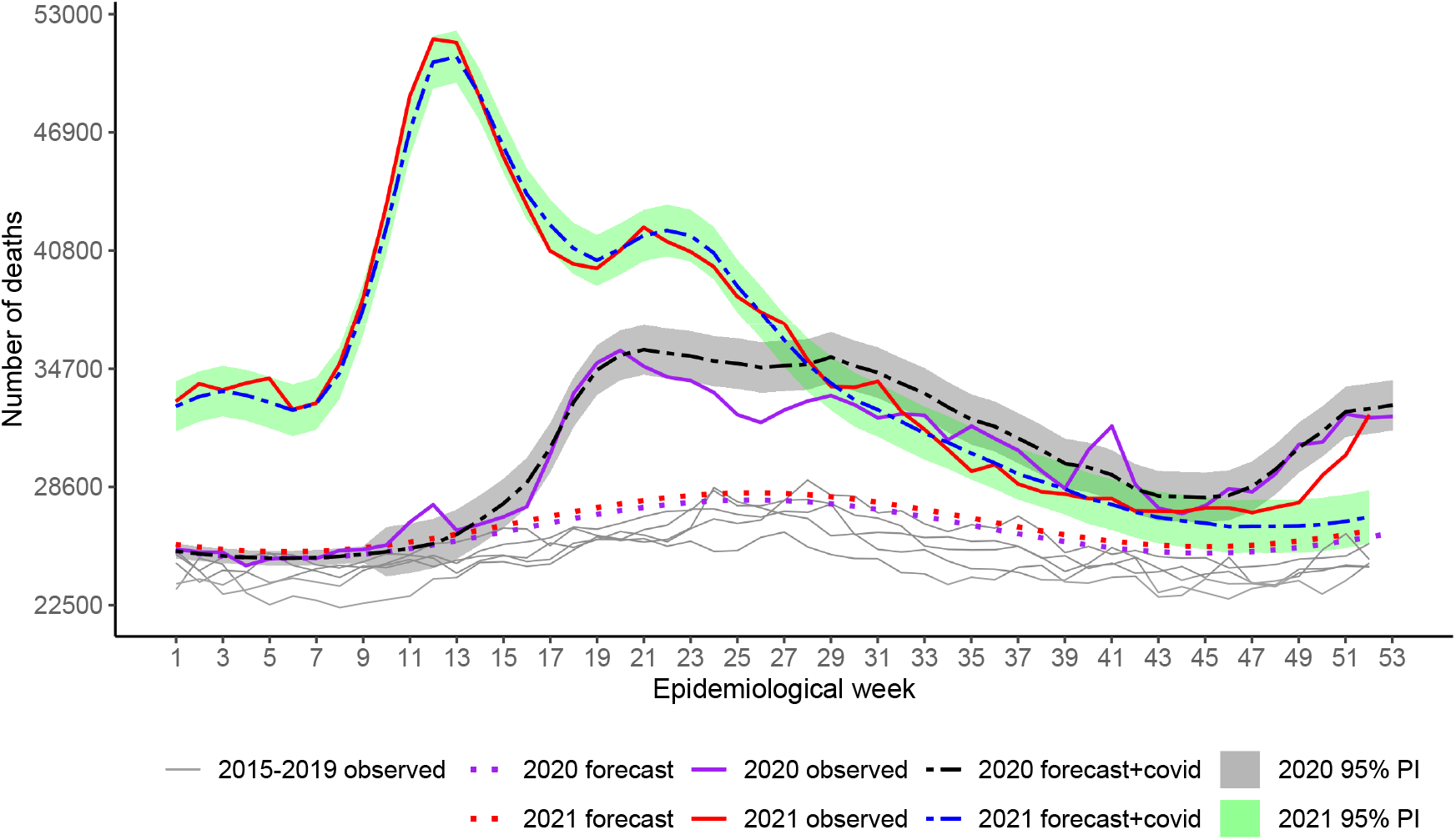
Weekly all-cause mortality in Brazil from week 1, 2015 to week 52, 2021. Recorded and baseline mortality forecast by the LMM and the forecast plus observed COVID-19 deaths including 95% prediction intervals (PI) for 2020 and 2021.

Excess has occurred over the whole period since the surge of the first wave in 2020 and reached alarming figures in the second wave in 2021. In the beginning, excess indicates that more deaths occurred beyond COVID-19. From week 13 to around week 22, 2020 (last week of June), excesses agreed well with COVID-19 as the primary cause. From week 22 to approximately week 32, 2020 (beginning of August), COVID-19 exceeded deaths from all causes, i.e., deaths due to other causes were smaller than expected. A peak of excess deaths occurred again in week 41 (around 10 October). From week 44 until the end of the year, deaths increased with the second wave, and excesses agree well with reported COVID-19 deaths. In 2021, the spread of variant Gamma before vaccination began, associated with further relaxation of social distancing, impacted mortality significantly. Only by mid of July 2021, death numbers lowered down to the level of the worst period of the previous year. For the occasion of the peak, around weeks 12-13 (end of March), only 2% of the country’s population was fully vaccinated. Vaccination for those classified as high-risk groups, e.g., elderly, and health workers, started very slowly on January 18, 2021 [41, 42]. As vaccination advanced, mortality tended to approach the expected figures, though, by December, excesses had increased again and reached the previous year’s levels.

Accumulated deaths in each year showed 214 620 COVID-19 and 187 842 estimated excess deaths (95% PI: 164 122 to 211 562; Ratio_EC_= 0.88) for weeks 10-53, 2020, and 420 193 COVID-19 and 441 048 estimated excess deaths (95% PI: 411 740 to 470 356; Ratio_EC_ = 1.05) for weeks 1-52, 2021 (Table 2). The P-Score estimates pointed out 16.1% and 31.9% more deaths than expected in the pandemic 2020 and 2021, respectively.

In Table 2, the summaries of expected and excess/deficit deaths accumulated according to specific causes and Fig 3 show that excesses and deficits occurred for most causes, at different points in time, except for Ill-defined Causes showing substantial deaths since the pandemic started, totaling 15 090 (23.5%) and 20 137 (25.0%) excesses in 2020 and 2021, respectively.

**Fig 3.**
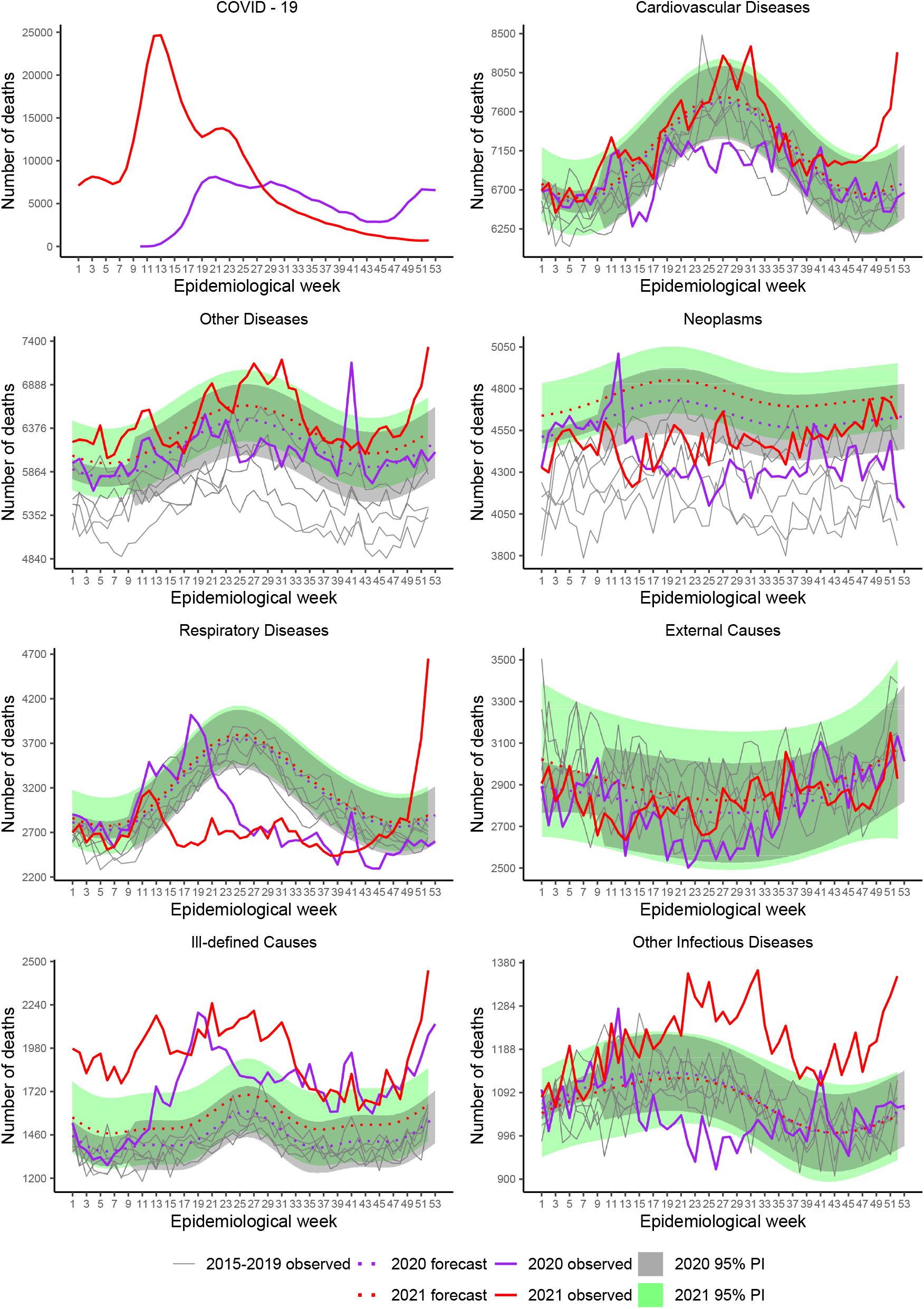
Weekly mortality in Brazil, stratified by death cause, from week 1, 2015 to week 52, 2021. Recorded and baseline mortality forecast by the LMM including 95% prediction intervals (PI) for 2020 and 2021.

For Respiratory, excesses occurred essentially at the beginning of the pandemic up to around week 20 (end of May 2020). After that, the impact was negative, with deficits until week 41 (early October) with the surge of a sudden spike. Overall, the effect was negative, showing a deficit of −14 980 (−10.6%). By week 12 (end of March) of 2021, we saw a peak, followed by decreasing figures lower than expected during almost the rest of the period, except that, by the end of November, there was a sharp increase. Overall, the balance was negative, showing a deficit of −22 291 (−13.5%). Mortality under Other Infectious Diseases showed a similar pattern to Respiratory in the year 2020, although on a smaller scale, with deficit deaths occurring during the weeks in the middle of the year (overall deficit of −1 797 and P-Score=−3.8%). However, in 2021, expressive mortality occurred, resembling the pattern of Ill-defined Causes (overall excess 7 087 and P-Score=12.7%).

Deficit deaths due to Cardiovascular Diseases occurred from March to August 2020, the period of tighter control measures. Overall the balance was −10 350 (−3.3%). In 2021, the mortality pattern followed the expected, except for the significant increase by the end of the year. Further analysis, when 2021 consolidated data are available, should be performed to confirm such an unexpected increase.

A peak stands out for Neoplasms at the beginning of the pandemic, followed by deficits for the rest of 2020. The year balance is −12 601 (−6.2%). The deficits persisted for most of the year 2021 with an overall balance of −13 466 (−5.5%). Note, however, that the baseline curves are, perhaps, overestimating, mainly for 2021. Such a pattern is due to the death series that showed a strong slope over time.

Overall, External Causes did not suffer the expected impact with the reduction of traffic and outdoor movements during restrictions, perhaps because many people were unable to comply with the recommendations.

From these inspections, we note that the first peak of excess deaths (week 12) in Fig 2 is related mainly to Ill-defined Causes, Neoplasms, Respiratory and Other Infectious Diseases. The peak at week 41, 2020, is related to these causes, except for Neoplasms.

### Excess deaths by state

Excess deaths occurred in all states, but with enormous heterogeneity across the country. Fig 4 and S1 Tab present the accumulated statistics for each state by the period. North and Central West regions states have P-Score values (left panel of 4) well above the value for Brazil (vertical lines), in both years, with Amazonas, Mato Grosso and the Federal District, with values ranging from 29.1-38.1% being the highlights in 2020. On the other hand, the states in the South are the highlights for the low percentages of excess death in that year, markedly Rio Grande do Sul with 1.4%. For 2021, the figures increased for all states, with Rondônia and Amazonas suffering alarming mortality, above 50%. Surprisingly, all states in the Northeast region have lower percentages than the global level, which, however, is quite large (31.9%).

**Fig 4.**
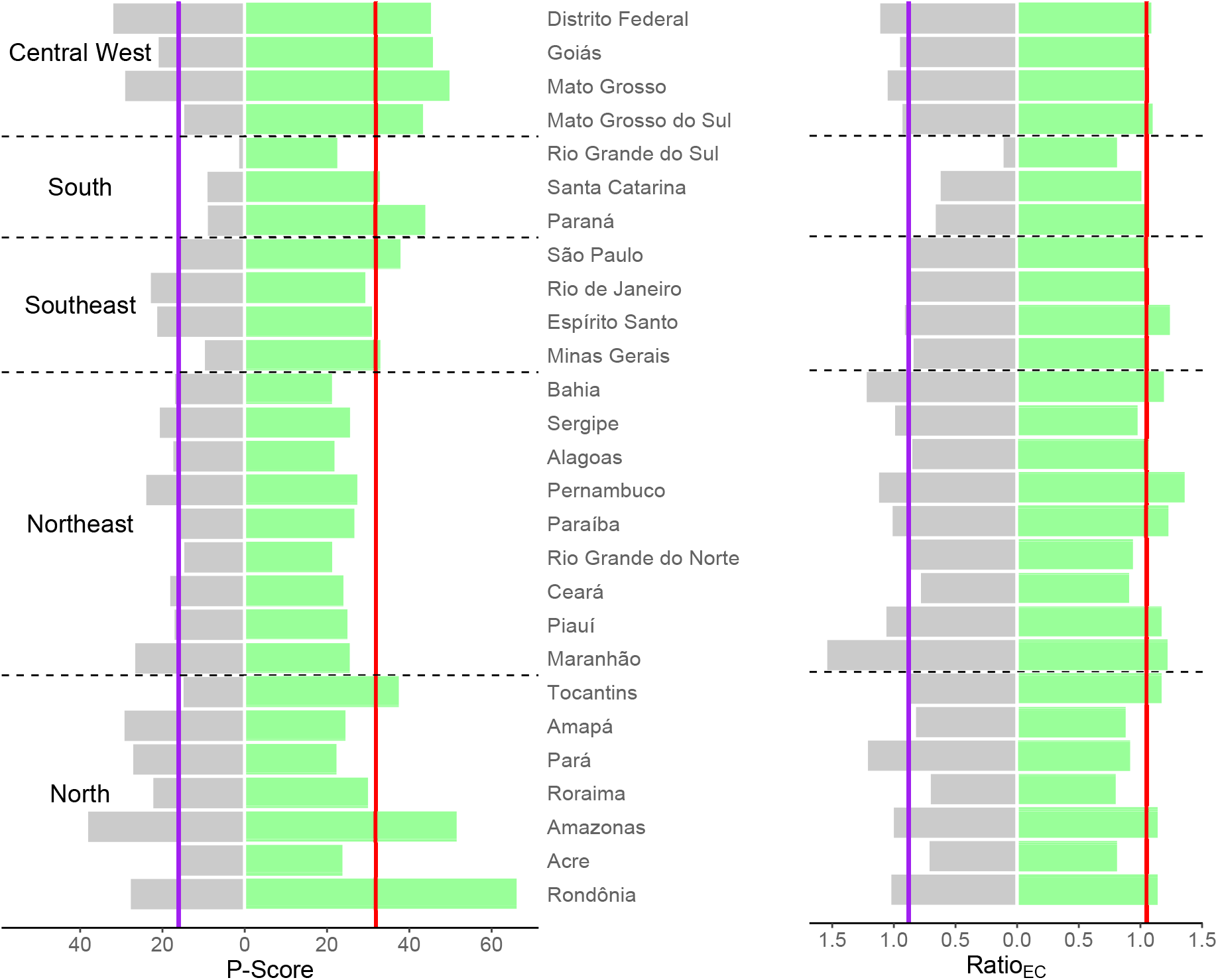
P-Score and Ratio_EC_ for each state, accumulated for two periods, weeks 10-53, 2020 (left in both panels) and weeks 1-52, 2021 (right in both panels). The vertical lines mark the values at the country level.

Several states suffered more excess deaths than COVID-19 in both years (right panel in Fig 4), mainly states in the Northeast, Central West and North regions. Deficit death was the rule, markedly in the South and Southeast regions, in the first year, with the lowest ratio (0.11) for Rio Grande do Sul. However, for the second wave, in 2021, most states showed ratios above 1, indicating that excess surpassed COVID-19 deaths. The minor figures were for Roraima and Acre in the North and Rio Grande do Sul in the South, all presenting ratios around 0.80.

Detailed results along time, shown in S2 Fig to S6 Fig, indicate that, in general, the states that coped better with the pandemic in the first year (as highlighted above) showed deficit deaths during the first wave, suggesting that their population afforded better care for other diseases and, perhaps, afforded better compliance to isolation recommendations. However, cause-diagnostic mistakes and COVID-19 under-reporting, mainly in remote states, could also contribute to the figures.

In 2021, deficit deaths are not clear in most of the states as the pandemic hit so badly the whole country and it was much more difficult for the population to practice isolation, except for Rio Grande do Sul, which shows a consistent pattern of deficit deaths. Still, some states showed notable positive discrepancies between forecast+COVID-19 deaths and reported deaths again, mainly in the North, Northeast and Central-West regions.

### Excess deaths according to sex, race/color and age

The SIM data is incomplete for sex, age and race/color, and so, before presenting the results on excess deaths according to these factors, we present their missing pattern. For sex and age, the percentages per year were small, just about 0.05% and 0.25% or less, respectively. Therefore, we do not expect biased results for these factors.

Fig 5 (A) and Table 3 show that for females, excess (13.4%) was smaller than for males (18.4%). Nonetheless COVID-19 surpassed excess during several weeks in the middle of 2020, producing Ratio_EC_= 0.76 for females, while for males, differences were smaller and for shorter period, resulting Ratio_EC_= 0.96. However, sex differences disappeared in 2021 with ratios of 1.03 and 1.07 and P-Score of 30.9% and 33.0% for females and males, respectively.

**Table 3.**
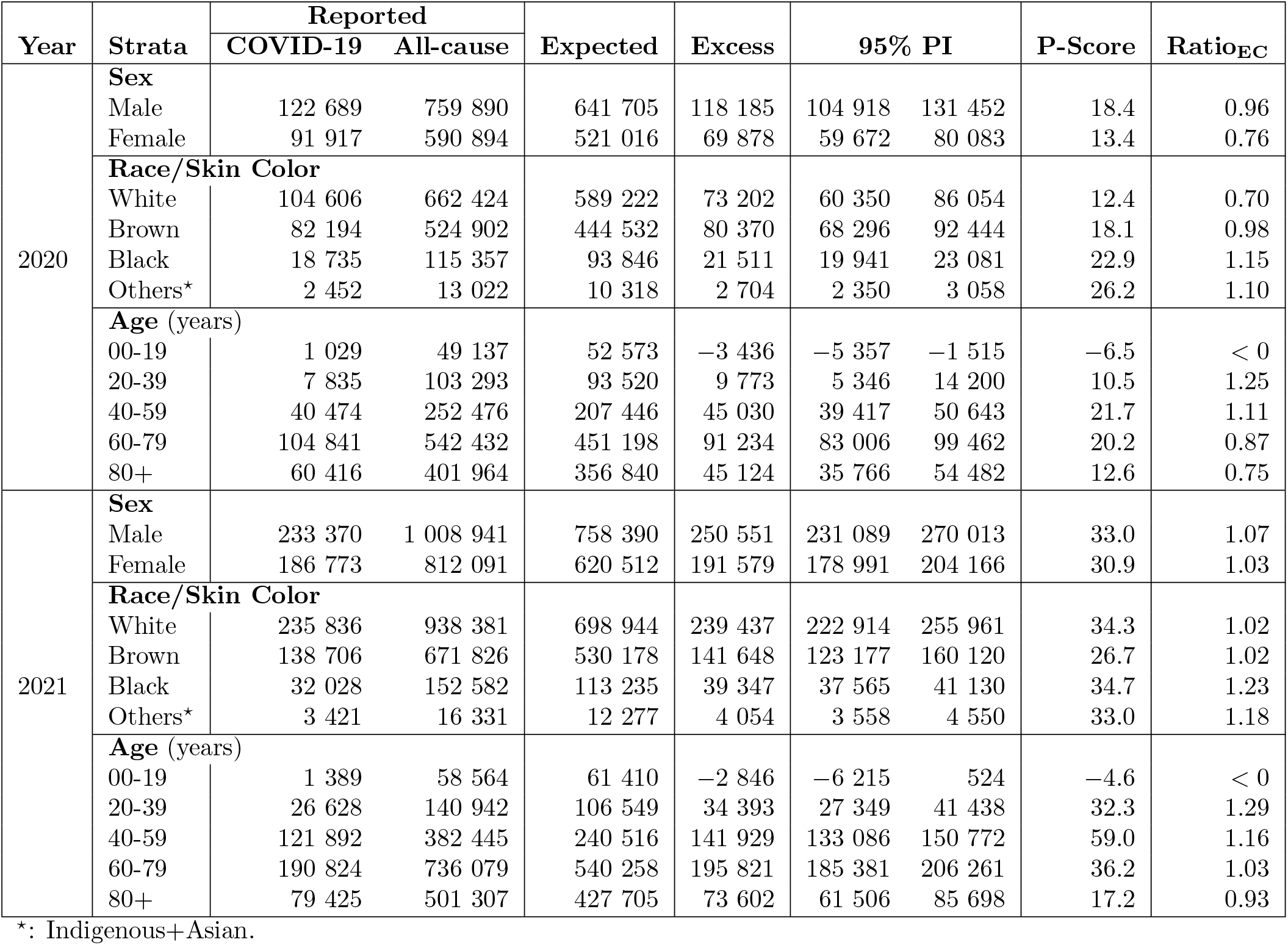
Reported, expected and estimated excess/deficit total deaths and deaths by sex, race/color and age group, accumulated for two periods, weeks 10-53, 2020 and weeks 1-52, 2021.

**Fig 5.**
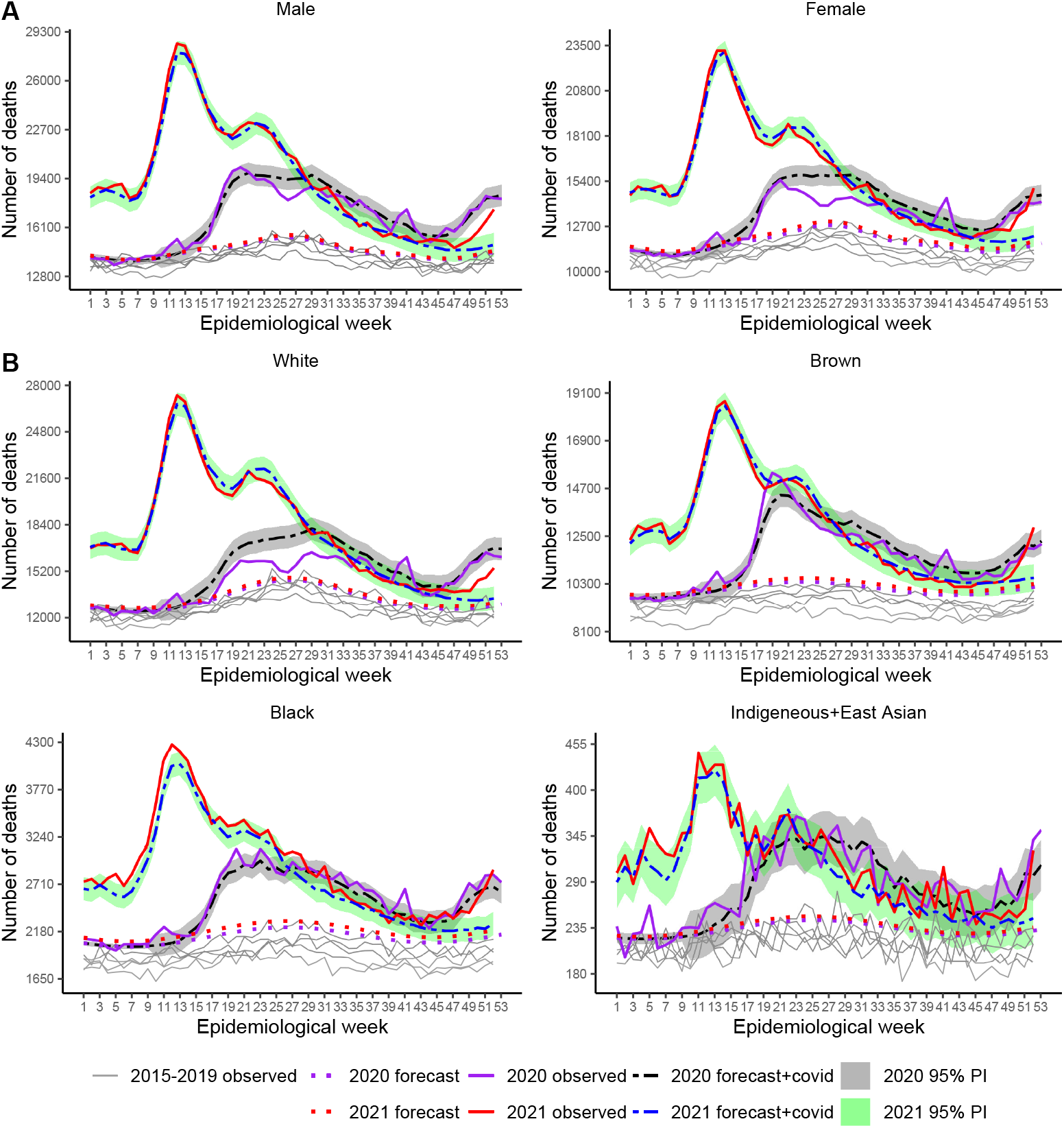
Weekly mortality in Brazil, stratified by sex (A) and race/color (B), from week 1, 2015 to week 52, 2021. Recorded and baseline mortality forecast by the LMM including 95% prediction intervals (PI) for 2020 and 2021.

The missingness for race/color appeared in outstanding percentages, ranging from 2.07 to 4.50%, with the earlier years presenting the larger shares. These figures are substantial since they exceed the *Others* category’s share (Indigenous or Asian ancestries, representing about 1.5% of the country population). The missing distribution across death-cause, sex, and age showed a homogeneous pattern so that we could consider it at random for these factors. However, across states, we found very high missing percentages in Alagoas (11.9 − 18.1%) and Bahia (4.1 − 9.9%), in the Northeast, and Espírito Santo (11.6 − 14.2%) and Minas Gerais (1.6 − 8.8%) in the Southeast region. While Bahia and Minas Gerais showed a clear decrease in the figures over the years, that was not the case for Alagoas and Espírito Santo. The problem seems more related to awareness of careful data recording than to places’ remoteness or availability of resources (or any other factor we could study). According to the last Brazilian census (2010), the shares of the joint categories Brown and Black to these state populations were 87% in Bahia, 67% in Alagoas, 57% in Espírito Santo and 53% in Minas Gerais (https://sidra.ibge.gov.br/Tabela/3145#resultado). For the country, the share of Black+Brown is about 50%. Consequently, we expect that these categories are under-represented in our analysis.

Fig 5 (B) shows deficit deaths for the White category only, during the first wave, when social distancing was recommended. For the Brown and Black categories, excess surpassed COVID-19 deaths at several periods, including 2021 for the Black. The Ratio_EC_ values ranged from 0.70 (White) to 1.15 (Black) in 2020, and from 1.02 (White) to 1.23 (Black) in 2021. The P-Score varied from 12.4% (White) to 26.2% (Others) and from 26.7% (Brown) to 34.7% (Black), in 2020 and 2021, respectively, showing how severely the black population was hit from both viewpoints, higher per capita mortality and excess deaths due to other causes beyond COVID-19.

Fig 6 shows the results stratified by age group. Deficit deaths prevailed in the group younger than 20 years old in 2020 (−6.5%) and 2021 (−4.5%). We note slight indication of deficit deaths, during the first wave, for the group 20-39 years old, but that disappeared from week 36, showing several peaks after September, 2020 and for 2021, so that the overall balance was positive, 10.5% in 2020 and 32.3% in 2021. This group presented the largest ratio, 1.25 in 2020 and 1.29 in 2021, indicating excess deaths due other causes than COVID-19. Note that, compared to the observed trajectories for the previous years, the baseline curves for 2020 and 2021 are lower. That is explained by a strong reduction tendency in the number of deaths, especially from 2016 to the weeks before the pandemic. Adults belonging to the 40-59 years old group show excess deaths surpassing COVID-19 practically over the two years, with ratio of 1.11 and 1.16 and P-Score of 21.7% and 50.9%, for 2020 and 2021, respectively. For the two oldest groups, the overall balance was positive as well, although there was a period of deficit deaths by the middle of the year 2020. The group of 80 years or older, experienced deficit deaths in 2021 as well, mainly after the start of vaccination, however that was canceled out by the step increase by the end of 2021. The ratios for this group were smaller than 1 in both years (0.75 and 0.93) indicating that, overall excess deaths had COVID-19 recorded as the main cause.

**Fig 6.**
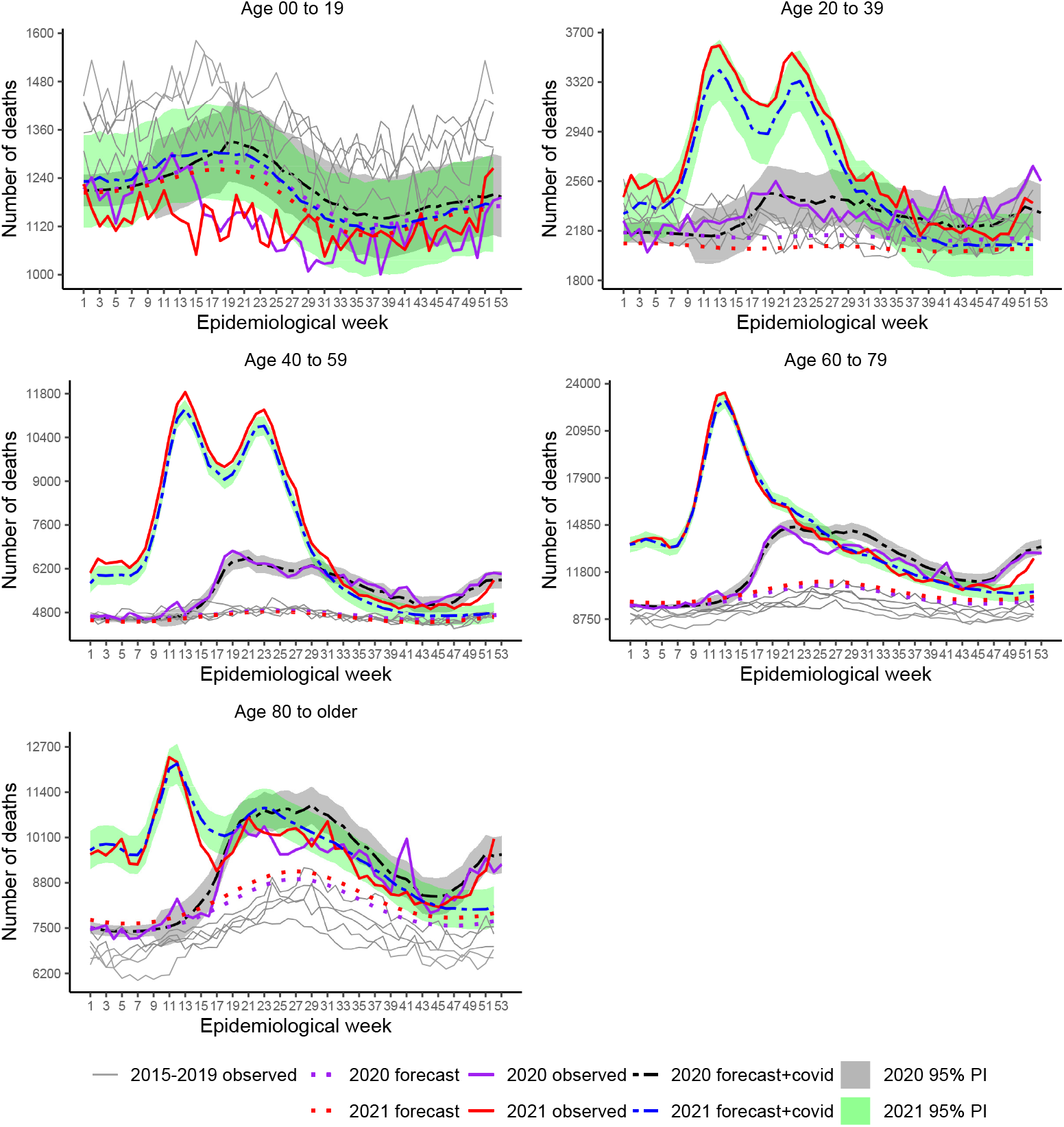
Weekly mortality in Brazil, stratified by age group, from week 1, 2015 to week 52, 2021. Recorded and baseline mortality forecast by the LMM including 95% prediction intervals (PI) for 2020 and 2021.

## Discussion

In this paper, we accessed the impact of the COVID-19 pandemic on Brazilian mortality over the years 2020 and 2021, by estimating excess deaths stratified by several factors.

Since the COVID-19 pandemic declaration in March 2020, excess deaths in Brazil amounted to 16.1% and 31.9% more than expected in 2020 and 2021, respectively. For 2020, our results unfold similar patterns published elsewhere (see [19] and [26], for example), although estimates vary because of the different modeling approaches, data updating, stratification factors and period considered. While we included only the pandemic weeks of 2020, other authors presented accumulated statistics for the entire year. Reported P-Score values are 13.7% ([19] using GLM assuming Negative Binomial distribution), 14% ([26] using an auto-regressive model, we guess, assuming normality), 19% ([43] using averaged previous five-year mortality rate) and 25% ([25] using averaged previous five-year death counts). As foreseen, using averaged statistics results in underestimated expected deaths and thus, in overestimated excess. As also reported in other studies [15, 16, 19], our investigation highlights the enormous heterogeneity across Brazil, showing that the South was less impacted (1.4-9.0%), while the North and Central-West had the highest P-Score values approaching 30% excess in several of them and almost 40% in the Amazonas, in 2020.

We are not aware of other publications that accessed 2021 excess, for as large a period as we have. [26] estimated 40% excess deaths up to week 14 of 2021, comprising the most critical period in terms of daily COVID-19 deaths in Brazil, when only about 8% of the population had at most the second vaccine dose [1]. That is larger than our estimate for the whole year (31.9%) but expected with the advance of vaccination since, by the end the year, 77% of the population was at least partially vaccinated. Across the country, our P-Score estimates ranged from around 20% (in four states, two in the Northeast and one in the South) to above 50% in two states in the North. While several factors resulting in societal inequalities are likely to contribute to regional differences, for Brazil, we should add the failure of the country to deliver, as a whole, the awareness of disease severity, the relevance of non-pharmacological measures and the benefits of vaccination. Looking at the states’ vaccination presently (end of 2022), for most of them, irrespective of the region, the share of at least one dose is above 75% [44], but with high variation within regions, namely, North (55-82%), Northeast (67-84%), Southeast (79-90%), South (83-87%) and Central West (73-89%).

Our results on deficit deaths from causes other than COVID-19 agree with those from other studies in which populations of more economically developed locations had a lower risk of death from other causes. Cause-specific expected mortality indicated deficits for Respiratory, Other Infectious Diseases, Neoplasms and Cardiovascular Diseases, in the year 2020, as also pointed out in [19], [26] and [43], although [26] and [43] used different death-cause grouping. With the social distancing recommendations, we expect lower exposure to risk factors associated with the first two, while deficits for Neoplasms and Cardiovascular diseases might be explained by the evidence that people suffering from these illnesses were at high risk for serious COVID-19 conditions and died from COVID-19 in the first wave [22, 26, 45–47]. While these arguments are plausible, we should keep in mind that misdiagnosis is always an issue, mainly in deaths that happened outside hospitals and clinics and in regions without the capacity for proper diagnosis [21]. Of these diseases, only Respiratory and Neoplasms maintained the deficit pattern in part of 2021.

Excesses deaths beyond COVID-19 were typical at the beginning of the pandemic, about the end of the first wave, and at the end of 2021, a signal of possible death cause misreporting [16, 19, 48, 49]. Excess deaths due to Ill-defined Causes occurred in both years with Other Infectious Diseases added in 2021 which show, as well, a steep increase of deaths from Respiratory and Cardiovascular diseases at the end of the year. Once definitive data for 2021 are available, it is crucial to reassess them to confirm or disregard such patterns.

Studies around the world [19, 22, 50–52] reported COVID-19 mortality affects more males than females, although some authors did not find relevant differences once baseline was considered [53]. Our results for 2020, based on excess deaths, are somewhat in line with [19], with 18.4% against 13.4% more extra deaths in males and females, respectively. However, for 2021, around 30% more deaths were estimated for both sexes. Our analysis pointed out more prominent female deficit deaths only during the plateau of the first wave, an indication that in 2020, women might have been able to engage better in social distancing and avoided contamination by other infections. That could be a consequence of the well-known vulnerable forms of employment (informal self-employing, housework and children care responsibilities) females share, mainly in South American countries [51].

The pandemic impacted age groups differently as expected and shown in other studies [19, 50, 52, 54] with a larger impact for people aged between 40 to 79 years old and 20 or older, in 2020 and 2021, respectively. The group younger than 20 showed deficit deaths in both years. In contrast, for the elderly, COVID-19 surpassed excess deaths, mainly in the first year, meaning fewer deaths from other causes. Some high-income countries also showed deficit death for youngsters [55]. Our deficit death estimate (−6.5%) is very close to that presented in [19] (−7.2%), and we believe their explanations are very plausible. Although the country did not apply strict lockdown, schools were closed, and outdoor and trip activities were reduced, contributing to lower exposure to external injuries and infections.

Our results showed excess exceeded deaths from COVID-19 in all race/color groups, except White, who had a deficit of deaths during part of the first wave. Contrasted to [19], our P-Score estimates are considerably larger for all groups except for the White but are lower than those presented in [13]. Nonetheless, all results go in the same direction that is, non-whites suffered higher excess deaths in the first year. [13] explored racial disparities for each region and showed the states contributing to marked disparities, with better figures for White, were those belonging to the South and Southeast, where the White population share is larger and, in general, enjoy better living conditions including access to prevention and healthcare assistance.

Strong points of our study are: our analyses, stratified by several factors, are based on the final data for 2020; we considered the most up-to-date 2021 data, including information for the whole year, the broader study we are aware of, and we used a flexible estimation method that allows explicit formulae for death forecast precision taking into account all uncertainty involved in the prediction process.

Our study has some limitations, data and estimates for 2021 are preliminary and expected to change due to the delay in reporting mortality. Health state secretaries have 60 days, following the end of the month of death occurrence, to fill out the national mortality system. After checking for inconsistencies and errors, a report is sent back for corrections, with the checking repeated on two or three occasions, causing a considerable delay in the availability of definitive data. As we followed 2020’s data from September 2021, when they were preliminary, until when they were declared definitive, we can say the differences were mainly related to death cause updating. The total mortality increased only 0.3% while COVID-19 death counts increased 1.5%. The most significant change was in mortality due to Ill-defined Diseases. We found that about 7% of the deaths preliminarily declared as Ill-defined Diseases migrated to other causes. Cardiovascular, Neoplasms and Other Diseases had, each, an increase of about 1%.

Another limitation is related to missing information in the data, in special for race/color. Our preliminary analysis revealed that the source of the missingness is related to a few states where Brown and Black groups’ share is larger than in the country’s overall population. That supports the argument that death counts for Brown and Black categories are under-represented, which can lead to under-estimated expected deaths. If the down-biases are proportional, the relative measures (P-Scores, Ratio_EC_) are not too seriously biased. There are alternatives to remedy the problem of missingness, such as using some missing imputation and bias-adjustment mechanisms. In the eminence of the availability of the 2022 census, some promising mechanisms might be devised.

## Supporting information

Step-by-step approach application

Technical methodological details

## Data Availability

Data are available from Ministerio da Saude, Governo do Brasil: https://opendatasus.saude.gov.br/dataset/sim-1979-2019 https://opendatasus.saude.gov.br/dataset/sim-2020-2021. However data from 2021 is considered preliminary and might be moved to another url once consolidated.

https://github.com/luziatrinca/COVID-19_excess_deaths_Brazil_2020_2021

## Supporting information

**S1 File. Technical methodological details**.

(PDF)

**S2 File. Application of the step-by-step approach for the baseline model**.

(PDF)

**S1 Tab. Reported, expected and estimated excess/deficit totals deaths by state, accumulated for two periods, weeks 10-53, 2020 and weeks 1-52, 2021**

**S1 Fig. Diagnostic graph analysis for assumptions of the fitted LMM for all-cause deaths in Brazil (Eq 4)**. Standardized marginal residuals against marginal fitted response, standardized conditional residuals against predicted response, Normal probability plot for standardized least confounded conditional residual, Chi-square probability plot for Mahalanobis distance and Modified Lesaffre-Verbeck measure index plot.

**S2 Fig. Weekly all-cause mortality in the North Region states, Brazil, from week 1, 2015 to week 52, 2021**. Recorded and baseline mortality forecast by the LMM and the forecast plus observed COVID-19 deaths including 95% prediction intervals for 2020 and 2021 (shaded areas).

**S3 Fig. Weekly all-cause mortality in the Northeast Region states, Brazil, from week 1, 2015 to week 52, 2021**. Recorded and baseline mortality forecast by the LMM and the forecast plus observed COVID-19 deaths including 95% prediction intervals for 2020 and 2021 (shaded areas).

**S4 Fig. Weekly all-cause mortality in the Southeast Region states, Brazil, from week 1, 2015 to week 52, 2021**. Recorded and baseline mortality forecast by the linear mixed model and the forecast plus observed COVID-19 deaths including 95% prediction intervals for 2020 and 2021.

**S5 Fig. Weekly all-cause mortality in the South Region states, Brazil, from week 1, 2015 to week 52, 2021**. Recorded and baseline mortality forecast by the linear mixed model and the forecast plus observed COVID-19 deaths including 95% prediction intervals for 2020 and 2021.

**S6 Fig. Weekly all-cause mortality in the Central-West Region states, Brazil, from week 1, 2015 to week 52, 2021**. Recorded and baseline mortality forecast by the linear mixed model and the forecast plus observed COVID-19 deaths including 95% prediction intervals for 2020 and 2021.

## References

1. Mathieu E, Ritchie H, Rodés-Guirao L, Appel C, Giattino C, Hasell J, et al. Coronavirus Pandemic (COVID-19); 2020. https://ourworldindata.org/coronavirus.

2. de Oliveira MM, Fuller TL, Gabaglia CR, Cambou MC, Brasil P, de Vasconcelos ZFM, et al. Repercussions of the COVID-19 pandemic on preventive health services in Brazil. Preventive Medicine. 2022;155. doi:10.1016/j.ypmed.2021.106914.

3. Malta M, Vettore MV, da Silva CMFP, Silva AB, Strathdee SA. Political neglect of COVID-19 and the public health consequences in Brazil: The high costs of science denial. EClinicalMedicine. 2021;35. doi:10.1016/j.eclinm.2021.100878.

4. Silva HM. The danger of denialism: lessons from the Brazilian pandemic. Bulletin of the National Research Centre. 2021;45. doi:10.1186/s42269-021-00516-y.

5. Ortega F, Orsini M. Governing COVID-19 without government in Brazil: Ignorance, neoliberal authoritarianism, and the collapse of public health leadership. Global Public Health. 2020; p. 1257–1277. doi:10.1080/17441692.2020.1795223.

6. da Silva LLS, Lima AFR, Polli DA, Razia PFS, Pavão LFA, Cavalcanti MAFDH, et al. Social distancing measures in the fight against COVID-19 in Brazil: Description and epidemiological analysis by state. Cad Saúde Pública. 2020;36:e00185020. doi:10.1590/0102-311X00185020.

7. Borrego-Morell JA, Huertas EJ, Torrado N. On the effect of COVID-19 pandemic in the excess of human mortality. The case of Brazil and Spain. PLoS ONE. 2021;16:1–14. doi:10.1371/journal.pone.0255909.

8. Barberia LG, Gómez EJ. Political and institutional perils of Brazil’s COVID-19 crisis. Lancet. 2020;396:367–368. doi:10.1016/S0140-6736(20)31681-0.

9. Burki T. No end in sight for the Brazilian COVID-19 crisis. Lancet Microbe. 2021;2:e180. doi:10.1016/S2666-5247(21)00095-1.

10. Azevedo e Silva G, Jardim BC, Santos CVB. Excesso de mortalidade no Brasil em tempos de COVID-19. Ciênc Saúde Colet. 2020;25:3345–3354. doi:10.1590/1413-81232020259.23642020.

11. da Fonseca EM, Nattrass N, Lazaro LLB, Bastos FI. Political discourse, denialism and leadership failure in Brazil’s response to COVID-19. Global Public Health. 2021;16:1251–1266. doi:10.1080/17441692.2021.1945123.

12. Brasil Ministério da Saúde. Banco de dados do Sistema Ùnico de Saúde-OpenDataSUS; 2022. https://opendatasus.saude.gov.br/dataset/sim-1979-2019; https://opendatasus.saude.gov.br/dataset/sim-2020-2021.

13. Teixeira RA, Vasconcelos AMN, Torens A, França EB, Ishitani L, Bierrenbach AL, et al. Excess Mortality due to natural causes among whites and blacks during the COVID-19 pandemic in Brazil. Revista da Sociedade Brasileira de Medicina Tropical. 2022;55. doi:10.1590/0037-8682-0283-2021.

14. Marinho MF, Torrens A, Teixeira R, Brant LCC, Malta DC, Nascimento BR, et al. Racial disparity in excess mortality in Brazil during COVID-19 times. Eur J Public Health. 2022;32:24–26. doi:10.1093/eurpub/ckab097.

15. Orellana JDY, Cunha GM, Marrero L, Moreira RI, da Costa Leite I, Horta BL. Excess deaths during the COVID-19 pandemic: underreporting and regional inequalities in Brazil. Cad Saúde Pública. 2021;37:1–16. doi:10.1590/0102-311X00259120.

16. Carvalho TA, Boschiero MN, Marson FAL. COVID-19 in Brazil: 150,000 deaths and the Brazilian underreporting. Diagn Microbiol Infect Dis. 2021;99:115258. doi:10.1016/j.diagmicrobio.2020.115258.

17. Helleringer S, Queiroz BL. Commentary: Measuring excess mortality due to the COVID-19 pandemic: progress and persistent challenges. Int J Epidemiol. 2022;51:85–87. doi:10.1093/ije/dyab260.

18. Verbeeck J, Faes C, Neyens T, Hens N, Verbeke G, Deboosere P, et al. A linear mixed model to estimate COVID-19-induced excess mortality. Biometrics. 2021;00:00–00. doi:10.1111/biom.13578.

19. Santos AM, Souza BF, Carvalho CA, Campos MAG, Oliveira BLCA, Diniz EM, et al. Excess deaths from all causes and by COVID-19 in Brazil in 2020. Rev Saúde Públ. 2021;55:1–12. doi:10.11606/s1518-8787.2021055004137.

20. Alves THE, Souza TA, Silva SA, Ramos NA, Oliveira SV. Underreporting of death by COVID-19 in Brazil’s second most populous state. Frontiers in Public Health. 2020;8:1–7. doi:10.3389/fpubh.2020.578645.

21. Brant LCC, Nascimento BR, Teixeira RA, Lopes MACQ, Malta DC, Oliveira GMM, et al. Excess of cardiovascular deaths during the COVID-19 pandemic in Brazilian capital cities. Heart. 2020;106:1898–1905. doi:10.1136/heartjnl-2020-317663.

22. Fernandes GA, Junior APN, Azevedo e Silva G, Feriani D, França e Silva ILA, Caruso P, et al. Excess mortality by specific causes of deaths in the city of São Paulo, Brazil, during the COVID-19 pandemic. PLoS ONE. 2021;16:e0252238. doi:10.1371/journal.pone.0252238.

23. Freitas ARR, Medeiros NM, Frutuoso LCV, Beckedorff OA, Martin LMA, Coelho MMM, et al. Tracking excess deaths associated with the COVID-19 epidemic as an epidemiological surveillance strategy-preliminary results of the evaluation of six Brazilian capitals. Rev Soc Bras Med Trop. 2020;53:e20200558. doi:10.1590/0037-8682-0558-2020.

24. Gimenez Junior GAA, Zilli PK, Silva LFF, Pasqualucci CA, Campo AB, Suemoto CK. Death trends based on autopsy data compared to the beginning of the coronavirus pandemic in Brazil. Braz J Med Biol Res. 2021;54:1–7. doi:10.1590/1414-431X202010766.

25. Lima EEC, Vilela EA, Peralta A, Rocha M, Queiroz BL, Gonzaga MR, et al. Investigating regional excess mortality during 2020 COVID-19 pandemic in selected Latin American countries. Genus. 2021;77. doi:10.1186/s41118-021-00139-1.

26. Nucci LB, Enes CC, Ferraz FR, da Silva IV, Rinaldi AEM, Conde WL. Excess mortality associated with COVID-19 in Brazil: 2020–2021. Journal of Public Health. 2021;doi:10.1093/pubmed/fdab398.

27. Marinho MF, Torrens A, Teixeira R, Brant LCC, Malta DC, Nascimento BR, et al. Racial disparity in excess mortality in Brazil during COVID-19 times. European Journal of Public Health. 2022;32:24–26. doi:10.1093/eurpub/ckab097.

28. IBGE-Instituto Brasileiro de Geografia e Estatística. Estudo Complementar à Aplicação da Técnica de Captura-Recaptura. Estimativas desagregadas dos totais de nascidos vivos e óbitos, 2016–2019; 2022. https://biblioteca.ibge.gov.br/visualizacao/livros/liv101927.pdf.

29. IBGE-Instituto Brasileiro de Estatística e Geografia. Desigualdades Sociais por Cor ou Raça no Brasil - Estudos e Pesquisas - Informação Demográfica e Socioeconômica; 2022. Available from: https://biblioteca.ibge.gov.br/visualizacao/livros/liv101972$_$informativo.pdf.

30. Karlinsky A, Kobak D. Tracking excess mortality across countries during the COVID-19 pandemic with the world mortality dataset. eLife. 2021;10:e69336. doi:10.7554/eLife.69336.

31. WHO-World Health Organization. Methods for estimating the excess mortality associated with the COVID-19 pandemic World Health Organisation; 2022. Available from: https://www.who.int/hac/about/.

32. Palacio-Mejía LS, andez Avila JEH, andez Avila MH, Dyer-Leal D, Barranco A, anchez ADQS, et al. Leading causes of excess mortality in Mexico duringthe COVID-19 pandemic 2020−2021: A deathcertificates study in a middle-income country. The Lancet Regional Health - Americas. 2022;13:100303. doi:10.1016/j.

33. Pinheiro JC, Bates D. Mixed-effects models in S and S-PLUS. New York: Springer-Verlag; 2000.

34. Singer JM, Rocha FMM, Nobre JS. Graphical tools for detecting departures from linear mixed model assumptions and some remedial measures. Int Stat Rev. 2017;85:290–324. doi:10.1111/insr.12178.

35. Aron J, Muellbauer J. The US excess mortality rate from COVID-19 is substantially worse than Europe’s; 2020. https://voxeu.org/article/us-excess-mortality-rate-covid-19-substantially-worse-europe-s.

36. R Core Team. R: A Language and Environment for Statistical Computing; 2021. Available from: https://www.R-project.org/.

37. Pinheiro J, Bates D, DebRoy S, Sarkar D, R Core Team. nlme: Linear and nonlinear mixed effects models; 2021. Available from: https://CRAN.R-project.org/package=nlme.

38. Baey C, Kuhn E. varTestnlme: variance components testing in mixed-effect models; 2019. Available from: https://github.com/baeyc/varTestnlme.

39. Halekoh U, Højsgaard S, Yan J. The R Package geepack for Generalized Estimating Equations. Journal of Statistical Software. 2006;15:1–11. doi:10.18637/jss.v015.i02.

40. Wickham H. ggplot2: Elegant Graphics for Data Analysis. Springer-Verlag New York; 2016. Available from: https://ggplot2.tidyverse.org.

41. Associação Médica Brasileira. Vacinação COVID-19 no Brasil: Passado, Presente e Desafios Futuros - AMB; 2021. https://amb.org.br/noticias/vacinacao-covid-19-no-brasil-passado-presente-e-desafios-futuros/.

42. Institute for Health Metrics and Evaluation - IHME. COVID-19; 2022. https://covid19.healthdata.org/brazil?view=cumulative-deaths&tab=trend.

43. Guimarães RM, de Oliveira MPRPB, Dutra VGP. Excess mortality according to group of causes in the first year of the COVID-19 pandemic in Brazil. Revista Brasileira de Epidemiologia. 2022;25. doi:10.1590/1980-549720220029.

44. Painel. Coronavírus Brasil; 2022. https://coronavirusbra1.github.io/.

45. Jardim B, Migowski A, Miranda Corrêa F, Azevedo e Silva G. Covid-19 in Brazil in 2020: impact on deaths from cancer and cardiovascular diseases. Revista de Saude Publica. 2022;56. doi:10.11606/S1518-8787.2022056004040.

46. Ng WH, Tipih T, Makoah NA, Vermeulen JG, Goedhals D, Sempa JB, et al. Comorbidities in SARS-CoV-2 patients: a systematic review and meta-analysis. mBio. 2021;12:1–12. doi:10.1128/mBio.03647-20.

47. Izcovich A, Ragusa MA, Tortosa F, Marzio MAL, Agnoletti C, Bengolea A, et al. Prognostic factors for severity and mortality in patients infected with COVID-19: A systematic review. PLoS ONE. 2020;15. doi:10.1371/JOURNAL.PONE.0241955.

48. Prado MF, Paula Antunes BB, Santos Lourenço Bastos L, Peres IT, Araújo Batista Silva A, Dantas LF, et al. Analysis of COVID-19 under-reporting in Brazil. Rev Bras Ter Intensiva. 2020;32:224–228. doi:10.5935/0103-507X.20200030.

49. Kupek E. How many more? Under-reporting of the COVID-19 deaths in Brazil in 2020. Trop Med Int Health. 2021;26:1019–1028. doi:10.1111/tmi.13628.

50. Cúellar L, Torres I, Romero-Severson E, Mahesh R, Ortega N, Pungitore S, et al. Excess deaths reveal the true spatial, temporal and demographic impact of COVID-19 on mortality in Ecuador. Int J Epidemiol. 2022;52:54–62. doi:10.1093/ije/dyab163.

51. PAHO Incident Management System Team and the PAHO Office for Equity, Gender and Cultural Diversity. COVID-19 Health Outcomes by Sex in the Americas. From January 2020 to January 2021; 2021. https://iris.paho.org/handle/10665.2/53372.

52. Aburto JM, Kashyap R, Schöley J, Angus C, Ermisch J, Mills MC, et al. Estimating the burden of the COVID-19 pandemic on mortality, life expectancy and lifespan inequality in England and Wales: a population-level analysis. J Epidemiol Community Health. 2021;75:735–740. doi:10.1136/JECH-2020-215505.

53. Krieger N, Chen JT, Waterman PD. Excess mortality in men and women in Massachusetts during the COVID-19 pandemic. Lancet. 2020;395:1829. doi:10.1016/S0140-6736(20)31234-4.

54. Michelozzi P, De’Donato F, Scortichini M, Pezzotti P, Stafoggia M, Sario MD, et al. Temporal dynamics in total excess mortality and COVID-19 deaths in Italian cities. BMC Public Health. 2020;20:1–8. doi:10.1186/S12889-020-09335-8.

55. Islam N, Shkolnikov VM, Acosta RJ, Klimkin I, Kawachi I, Irizarry RA, et al. Excess deaths associated with covid-19 pandemic in 2020: Age and sex disaggregated time series analysis in 29 high income countries. The BMJ. 2021;373. doi:10.1136/bmj.n1137.

