## Supplementary material for "Assessing COVID-19 pandemic excess deaths in Brazil: years 2020 and 2021": Step-by-step approach application

### Application of the step-by-step approach for the baseline model

#### Excess deaths at the country level

In this supporting information we present detailed results for the application of the step-by-step strategy for selecting a baseline model for mortality,described in the paper. We use death from all-cause at the country level as an illustration.

Table 1 shows the fit information for modeling the all-cause weekly deaths at the country level. Each row shows the model fitted in each step of the strategy previously outlined. Two terms of the Fourier series (Model 1) with the estimated frequency $\hat{\omega} = 0.1598 \approx 0.16$, resulting in a period of approximately 39 weeks, showed a good approximation for the fixed part (lack-of-fit p-value= 0.6322 when comparing the simpler model to the full Model 0). The first fitted mixed model (Model 2) included six fixed parameters (the intercept plus four

Table 1: **Baseline model fitting information for all-cause weekly deaths based on historical data from week 1, 2015 to week nine, 2020**

| **Model** | **Method** | **Error structure** | **Number of parameters** | **Global fit**$^a$ | **p-value**$^b$ |
|---|---|---|---|---|---|
| 0 | OLS | $\sigma^2\mathbf{I}$ | 57 | 74 540 015 | |
| 1 | OLS | $\sigma^2\mathbf{I}$ | 11 | 89 319 620 | 0.6322 |
| 2 | REML | $\sigma^2\mathbf{I}$ | 22 | −2 043.990 | |
| 3 | REML | $\sigma^2\mathbf{I}$ | 12 | −2 050.965 | 0.1752 |
| 4 | REML | $\sigma^2\mathbf{I}$ | 11 | −2 050.964 | 0.9999 |
| 5 | REML | AR(1) | 12 | −2 000.257 | $< 0.0001$ |
| 6 | REML | AR(1) | 11 | −1 999.993 | 0.4674 |
| 7 | ML | AR(1) | 11 | −2 034.261 | |
| 8 | ML | AR(1) | 9 | −2 034.852 | 0.5537 |
| 9 | REML | AR(1) | 9 | −2 011.445 | |

*a*: global fit measures are Residual Sum of Squares (OLS) and log of the likelihood (REML/ML).
*b*: lack-of-fit test comparing the current model to the previous one.

slopes for two waves of the FS and the linear effect of time) and five random components, with unstructured $\mathbf{D}$ and diagonal $\mathbf{R} = \sigma^2\mathbf{I}$ (no serial correlations and homogeneous errors), such that the total number of the model parameters was 22 ($p = 5$ fixed terms and $q_{\boldsymbol{\theta}} = 5 + 10 + 1 = 16$ associated with the random effects).

The sequence of steps shows simplifications that could be applied and improvements brought by the inclusion of AR(1) structure for serial correlation. Models 7, 8 and 9 were generated in the last steps of the strategy described in the paper. Note the requirement to change the estimation method for simplification of the mean model (fixed part) as explained in the S1 File.
