## Supplementary figures and images for "Assessing COVID-19 pandemic excess deaths in Brazil: years 2020 and 2021"

### S1 Fig

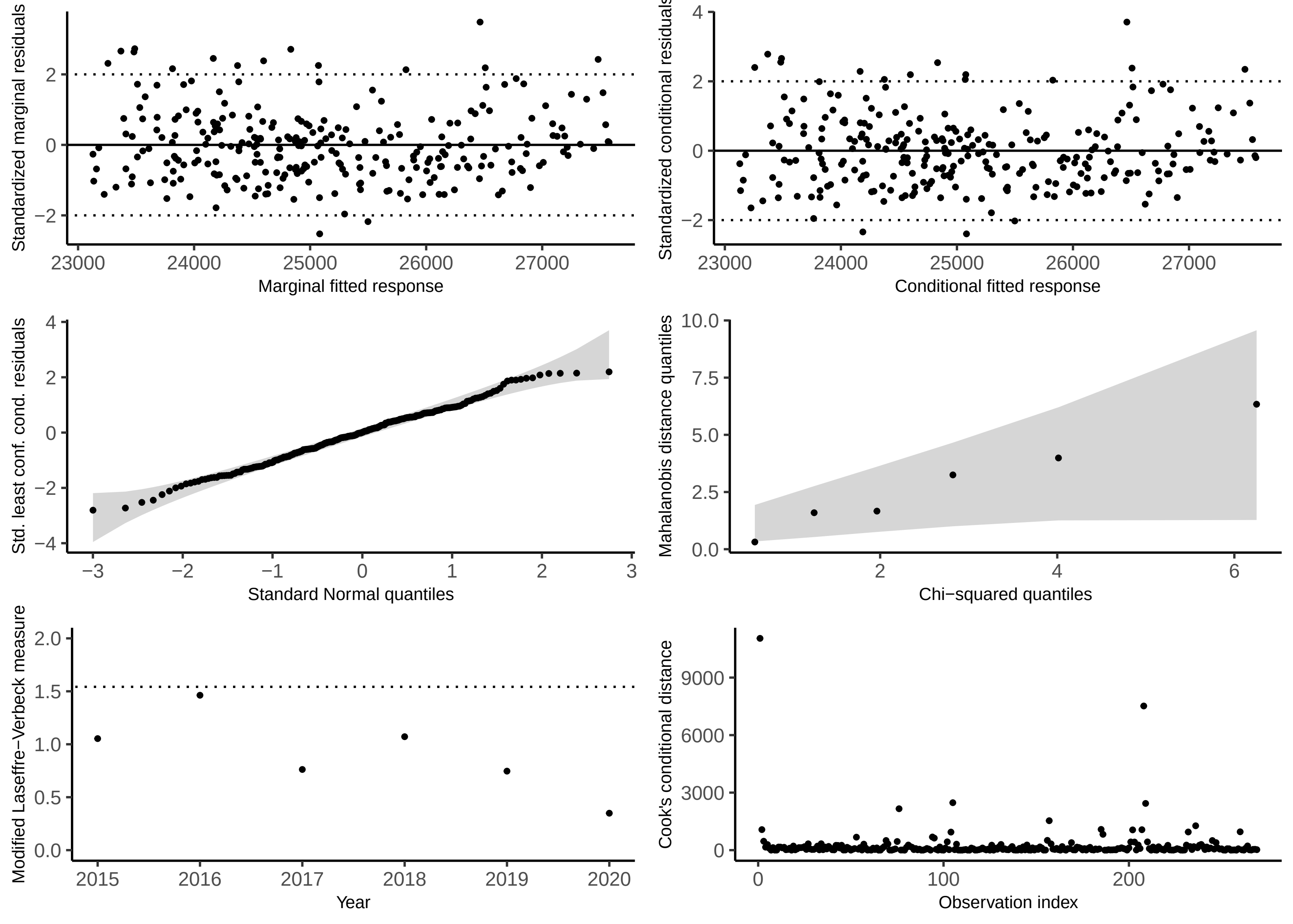

### S2 Fig

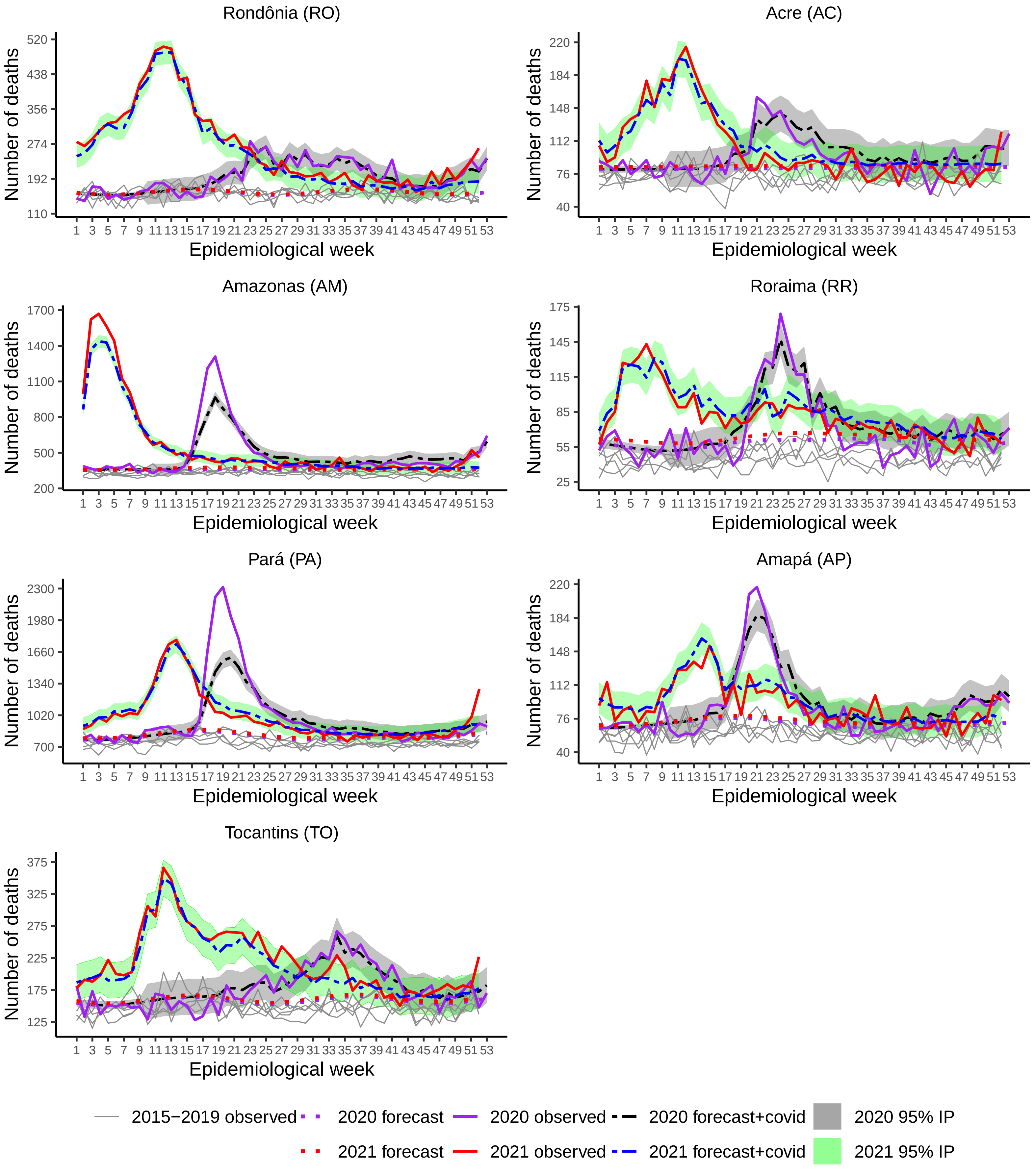

### S3 Fig

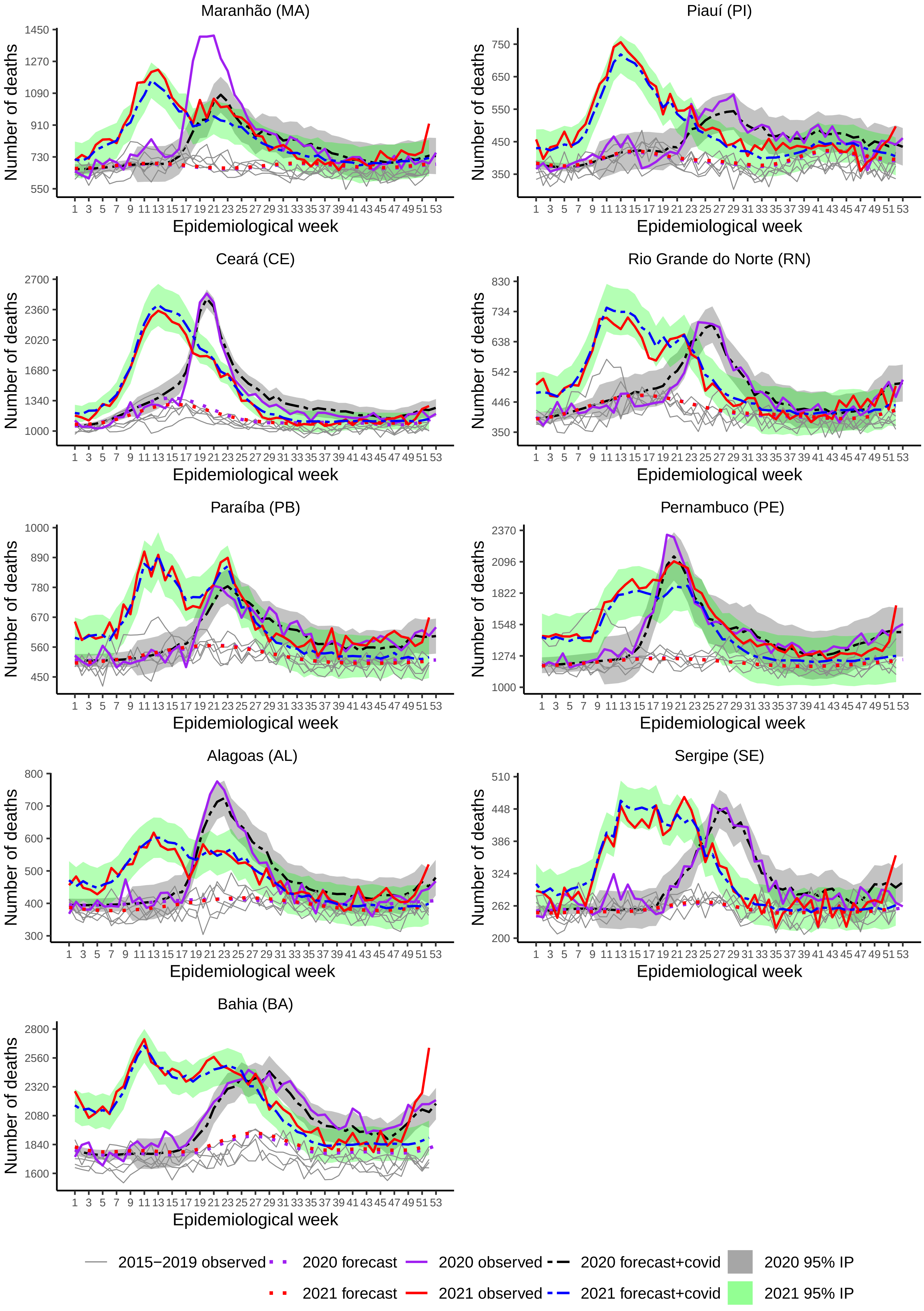

### S4 Fig

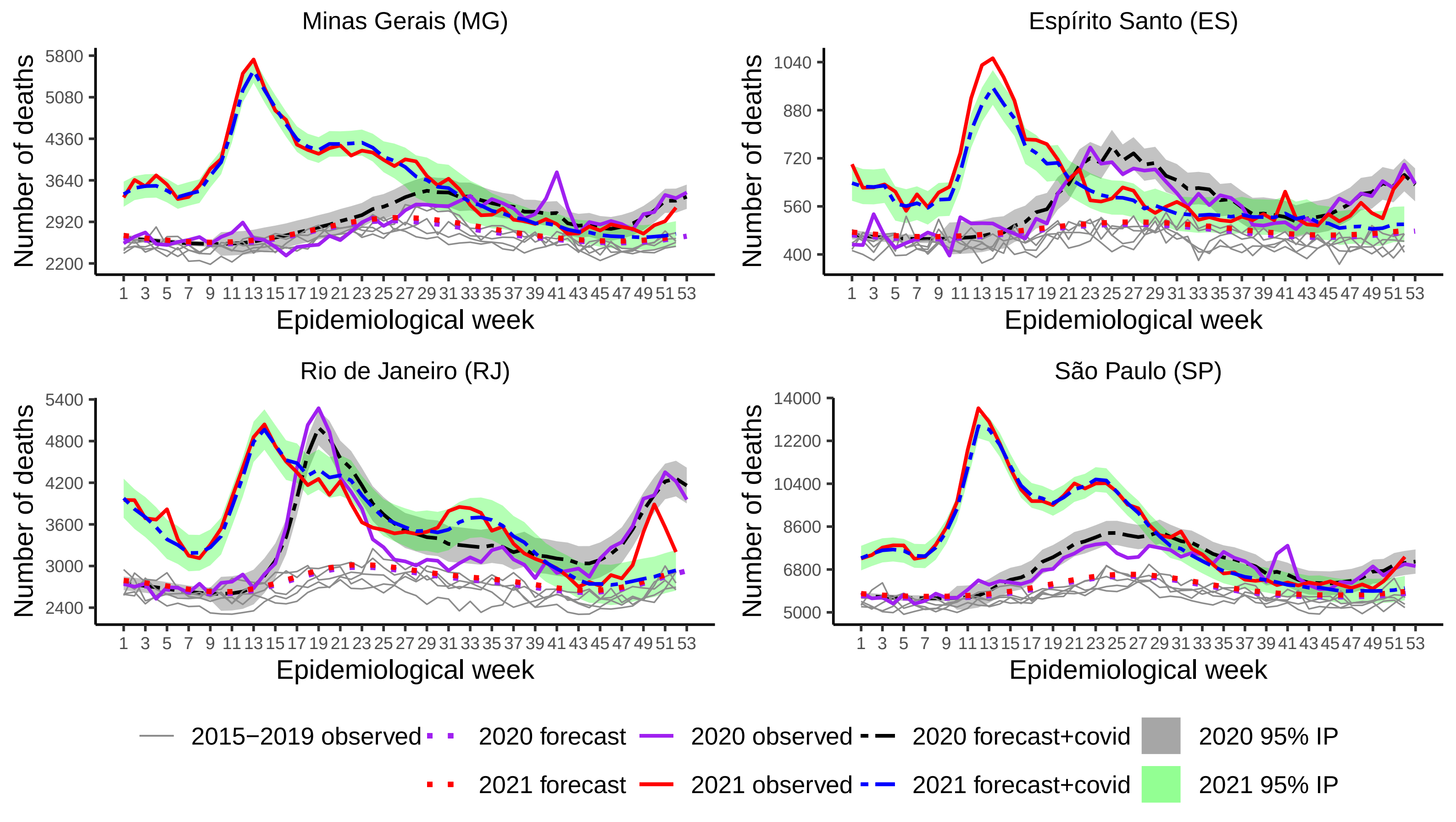

### S5 Fig

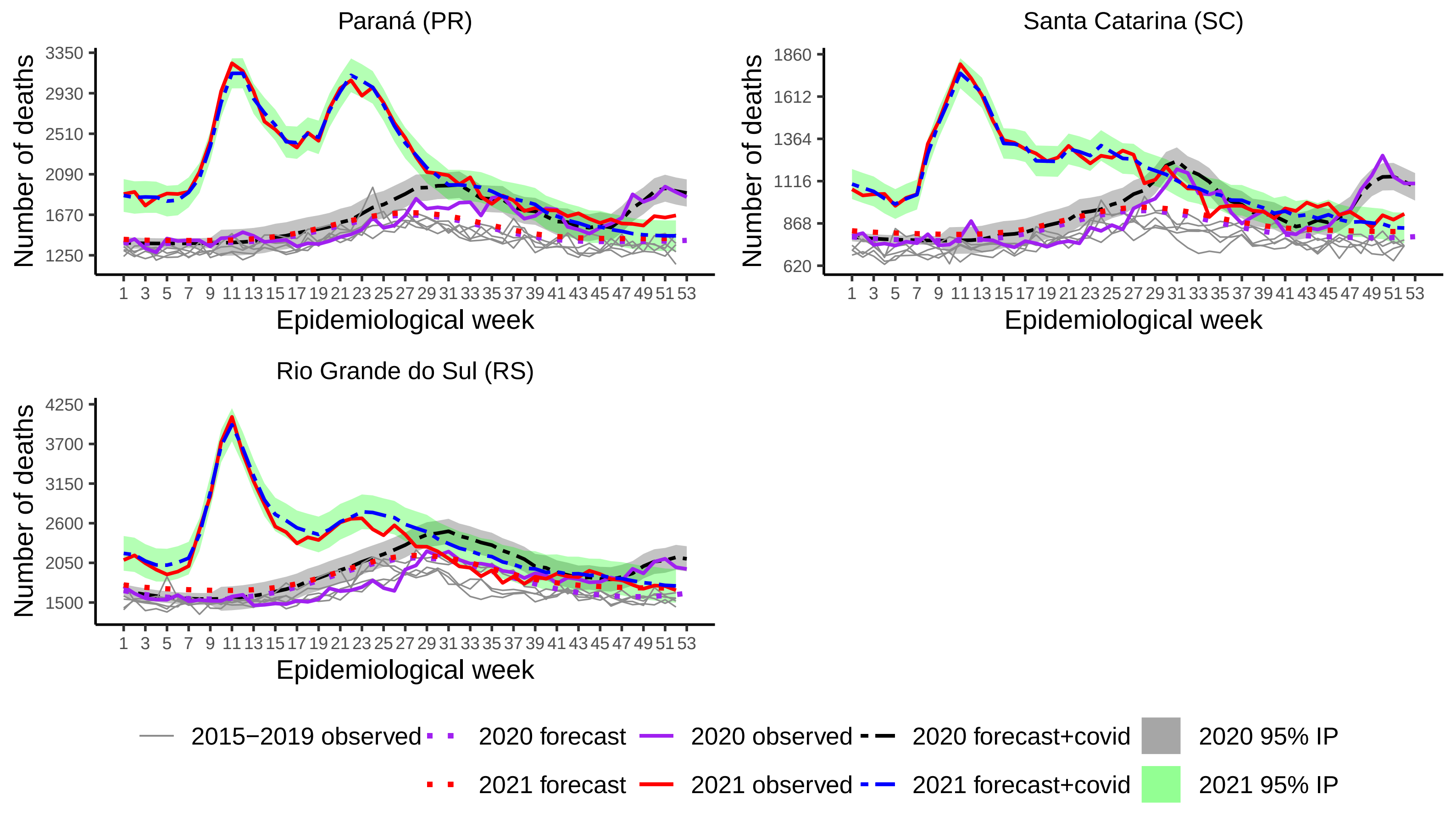

### S6 Fig

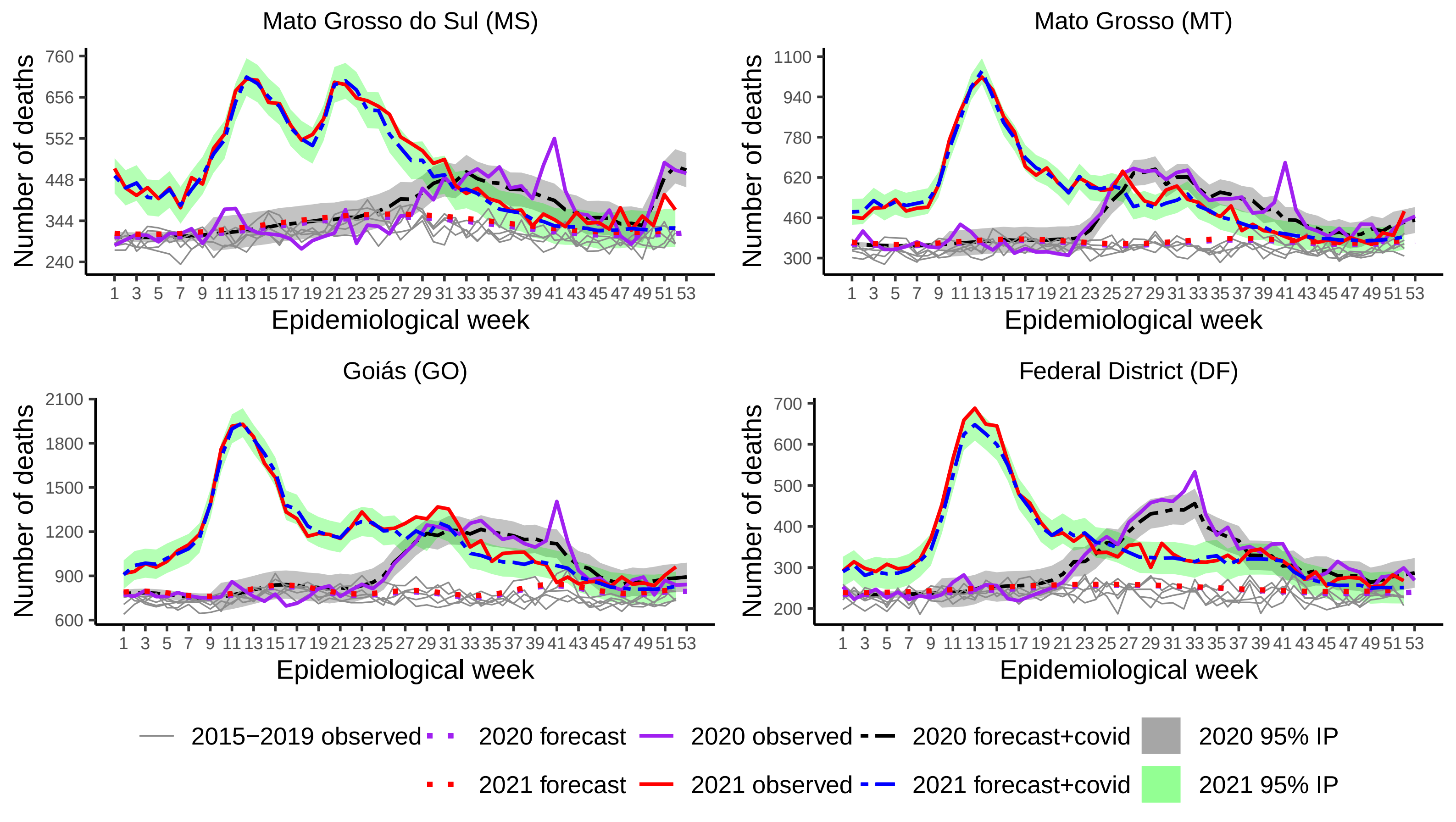
