## Supplementary material for "Assessing COVID-19 pandemic excess deaths in Brazil: years 2020 and 2021": Technical methodological details

Saditt Rocio Robles Colonia[1], Lara Morena Cardeal[1], Rogério Antonio de Oliveira[2], Luzia Aparecida Trinca[2]
**1** Research Program in Biometry, Unesp, Botucatu, São Paulo, Brazil
**2** Institute of Biosciences, Unesp, Botucatu, São Paulo, Brazil

### Technical methodological details

#### The linear mixed model

In the mixed model framework, each year is considered a unit or cluster and weeks within a year are the observational units. A linear mixed model involves the modeling of three components: the mean or fixed-effects structure, the random-effects structure and the serial correlation structure. Using standard notation, let $\mathbf{Y}_i$ denote the response vector ($T_i \times 1$) for year $i$ ($i = 1,\ 2,\ \ldots, M$). $T_i$ refers to the number of epidemiological weeks of year $i$ and $M$ is the number of years in the historical data ($M = 6$ for years 2015-2020). For the period considered, all years have 52 weeks except 2020 that has 53 weeks. However, for the mixed model, 2020 data from weeks 10 to 53 are considered missing since this is the pandemic period and the mixed model will be used to predict baseline mortality under non pandemic circumstances. The LMM is

$$\mathbf{Y}_i = \mathbf{X}_i\boldsymbol{\beta} + \mathbf{Z}_i\mathbf{b}_i + \boldsymbol{\varepsilon}_i, \tag{1}$$

wherein $\mathbf{X}_i$ is the mean model matrix ($T_i \times p$); $\boldsymbol{\beta}$ is the vector of fixed parameters ($p \times 1$); $\mathbf{Z}_i$ is the model matrix for the random terms ($T_i \times q$); $\mathbf{b}_i \sim N_q(\mathbf{0};\ \mathbf{D}_i)$ is the year-specific random effects vector and $\boldsymbol{\varepsilon}_i \sim N_{T_i}(\mathbf{0};\ \mathbf{R}_i)$ is the random measurement errors vector. As standard, $\mathbf{b}_i$ and $\boldsymbol{\varepsilon}_i$ are assumed independent. There are many choices for each part of the model. The columns of $\mathbf{X}_i$ could be indicators for weeks or could accommodate some function of weeks, as a quantitative explanatory variable, selected from a wide class of functions. The selected function should capture seasonal and trend effects. Other relevant covariates for explaining the mean of death counts could be included as well. The columns of $\mathbf{Z}_i$ depend on the required random components to explain clustering and variability from year-to-year death patterns. The forms of the covariance matrices $\mathbf{D}_i$ and $\mathbf{R}_i$ should account for the variances and correlation patterns of the random terms and depend on other parameters $\boldsymbol{\theta}_i$. Usually, it is assumed the random components share the same distribution among units, that is, $\boldsymbol{\theta}_i = \boldsymbol{\theta}$, $\mathbf{D}_i = \mathbf{D}$ and $\mathbf{R}_i = \mathbf{R}$ for $i = 1, 2, \ldots, 6$, but that is not a requirement. In that case, $\boldsymbol{\theta}$ is a $q_{\boldsymbol{\theta}} \times 1$ vector. For the model under these assumptions and further independence among all random terms, that is, $\mathbf{D}$ and $\mathbf{R} = \sigma^2\mathbf{I}$ diagonal matrices, $q_{\boldsymbol{\theta}} = q + 1$. The model in (1) implies that

$$\mathrm{E}(\mathbf{Y}_i) = \mathbf{X}_i\boldsymbol{\beta} \ \text{ and } \ \mathrm{V}(\mathbf{Y}_i) = \mathbf{Z}_i\mathbf{D}\mathbf{Z}_i{}^\top + \mathbf{R} = \boldsymbol{\Sigma}_i(\boldsymbol{\theta}) \tag{2}$$

the so called marginal model ($\mathbf{Z}_i{}^\top$ denotes the transposed $\mathbf{Z}_i$). Given the year-specific effects $\mathbf{b}_i$, we have the conditional model,

$$\mathrm{E}(\mathbf{Y}_i|\mathbf{b}_i) = \mathbf{X}_i\boldsymbol{\beta} + \mathbf{Z}_i\mathbf{b}_i \ \text{ and } \ \mathrm{V}(\mathbf{Y}_i|\mathbf{b}_i) = \mathbf{R} \tag{3}$$

responsible for predicting year-specific baseline mortality once the parameters are estimated. The bar "|" refers to the concept of conditional random variables, in this case mortality vector for year $i$ conditioned on year $i$ specific effects $\mathbf{b}_i$.

Classical methods for estimation of the LMM parameters are Maximum Likelihood (ML) and Restricted Maximum Likelihood (REML) [**?**, **?**, **?**]. It is known that, given $\boldsymbol{\theta}$, the $\boldsymbol{\beta}$ ML estimator can be obtained from the generalized least squares formula (mixed model equations),

$$\widehat{\boldsymbol{\beta}} = \left(\sum_{i=1}^{M}\mathbf{X}_i^\top(\boldsymbol{\Sigma}_i(\boldsymbol{\theta}))^{-1}\mathbf{X}_i\right)^{-1}\sum_{i=1}^{M}\mathbf{X}_i^\top(\boldsymbol{\Sigma}_i(\boldsymbol{\theta}))^{-1}\mathbf{Y}_i \tag{4}$$

which is the *best linear unbiased estimator* (BLUE) for $\boldsymbol{\beta}$. Predictions for the random effects are also derived

$$\tilde{\mathbf{b}}_i = \mathbf{D}\mathbf{Z}_i{}^\top(\boldsymbol{\Sigma}_i(\boldsymbol{\theta}))^{-1}(\mathbf{Y}_i - \mathbf{X}_i\widehat{\boldsymbol{\beta}}) \tag{5}$$

resulting in the *best linear unbiased predictor* (BLUP) for $\mathbf{b}_i$. Note that $\widehat{}$ is used to imply the function is an *estimator* of fixed parameters (population constants), while $\tilde{}$ is used to refer the function is a *predictor* of a latent variable.

For unknown $\boldsymbol{\theta}$, both parameter vectors can be estimated by ML using some iterative procedure, however, rendering biased estimates for $\boldsymbol{\theta}$. REML, proposed by [?], is preferable since it accounts for the degrees of freedom loss in the process of $\boldsymbol{\beta}$ estimation and is expected to yield less biased estimators for $\boldsymbol{\theta}$. Substituting $\hat{\boldsymbol{\theta}}$ in Eqs (4) and (5) results in the so called empirical BLUE and empirical BLUP, respectively.

#### Baseline death prediction intervals

Let $\widehat{\boldsymbol{\beta}}$ be the empirical BLUE of $\boldsymbol{\beta}$ (Eq 4) and $\tilde{\mathbf{b}}_6$ be the empirical BLUP of $\mathbf{b}_6$ (Eq (5)$i = 6$ refers to 2020). The 2020-specific conditional empirical best linear unbiased predictor from the final model was used as the 2020 baseline mortality. It is instructive to write the 2020 prediction vector in two parts, one for weeks 1-9 (pre-pandemic, data is available and used to fit the model), given by

$$\tilde{\mathbf{y}}_6^{pre}|(\mathbf{b}_6 = \tilde{\mathbf{b}}_6) = \mathbf{X}_{pre}\widehat{\boldsymbol{\beta}} + \mathbf{Z}_{pre}\tilde{\mathbf{b}}_6, \tag{6}$$

wherein *pre* stands for the part of the matrices referring to the pre-pandemic period. For weeks 10-53 (pandemic period, no data are available) the predictions are

$$\tilde{\mathbf{y}}_6^{pan}|(\mathbf{b}_6 = \tilde{\mathbf{b}}_6) = \mathbf{X}_{pan}\widehat{\boldsymbol{\beta}} + \mathbf{Z}_{pan}\tilde{\mathbf{b}}_6 + \boldsymbol{\varepsilon}_{6pan}, \tag{7}$$

wherein *pan* refers to the pandemic period. For obtaining $\tilde{\mathbf{y}}_6^{pan}|\tilde{\mathbf{b}}_6$ in (7) we substitute $\boldsymbol{\varepsilon}_{6pan}$ by its best predictor available, i.e the null vector, and the distinction between predictions formulae (6) and (7) is with respect to their variances. The variance-covariance matrix of the complete predicted vector $\tilde{\mathbf{y}}_6|\mathbf{b}_6$ is

$$\widehat{\mathrm{V}}(\tilde{\mathbf{y}}_6|\mathbf{b}_6) = \begin{pmatrix} \mathbf{X}_{pre}\widehat{\mathrm{V}}(\widehat{\boldsymbol{\beta}})\mathbf{X}_{pre}^\top \\ \mathbf{X}_{pan}\widehat{\mathrm{V}}(\widehat{\boldsymbol{\beta}})\mathbf{X}_{pan}^\top + \widehat{\mathbf{R}}_{pan} \end{pmatrix}, \tag{8}$$

wherein $\widehat{\mathbf{R}}_{pan}$ is the block of the matrix $\widehat{\mathbf{R}}$ with respect to weeks 10-53 and $\widehat{\mathrm{V}}(\widehat{\boldsymbol{\beta}}) = \left(\sum_{i=1}^{M}\mathbf{X}_i^\top(\boldsymbol{\Sigma}_i(\hat{\boldsymbol{\theta}}))^{-1}\mathbf{X}_i\right)^{-1}$ is the first-order approximation estimator of the variance-covariance of $\widehat{\boldsymbol{\beta}}$. Note that for the pandemic period, we want to forecast non-observable mortality and variability due to the random measurement error should be taken into account. The fitted model offers a forecast for week 53 since this time point is close to the upper limit of the time range of the data used for the baseline modeling, e. g. extrapolation error is not a big issue here. We need to accommodate $\widehat{\mathbf{R}}_{pan}$ as well for week 53. Under the homogeneous errors assumption, which we will see the data show no violation, this is easy do deal with.

For the year 2021 ($i = 7$), no non-pandemic data is observable and there is no prediction of the year-specific effect. Thus, $\tilde{\mathbf{y}}_7|\mathbf{b}_7$ is

$$\tilde{\mathbf{y}}_7|\mathbf{b}_7 = \mathbf{X}\widehat{\boldsymbol{\beta}} + \mathbf{Z}\mathbf{b}_7 + \boldsymbol{\varepsilon}_7, \tag{9}$$

and the best predictors for both, $\mathbf{b}_7$ and $\boldsymbol{\varepsilon}_7$, are zero, from the assumed model. Then, the variance-covariance is

$$\widehat{\mathrm{V}}(\tilde{\mathbf{y}}_7|\mathbf{b}_7) = \mathbf{X}\widehat{\mathrm{V}}(\widehat{\boldsymbol{\beta}})\mathbf{X}^\top + \mathbf{Z}\widehat{\mathbf{D}}\mathbf{Z}^\top + \widehat{\mathbf{R}}. \tag{10}$$

For prediction intervals (PI), standard errors ($\widehat{se}_{it}$) are estimated by the squared root of the diagonal elements of (8) and (10). Approximate 95% PI's are calculated using the standard method (Normal approximation):

$$PI(y_{it}|\mathbf{b}_i;\ 95\%) = \left[\tilde{y}_{it}|\mathbf{b}_i - 2\times\widehat{se}_{it};\ \tilde{y}_{it}|\mathbf{b}_i + 2\times\widehat{se}_{it}\right]. \tag{11}$$

#### Modeling by GEE

#### Prediction Accuracy

To assess the prediction accuracy of the LMM we used the square root of forecast error (RMSE) and the mean absolute percentage error (MAPE). For this task, we used data from 2010-2019 such that for each year $i$ in turn, from 2015 to 2019 ($i = 1,\ 2,\ \ldots,\ 5$), we fitted each model being compared using data from the previous five-year history and the 9 first weeks of year $i$. Then, for year $i$, we obtained forecasts for $t = 10,\ 11,\ \ldots,\ 52$ and evaluated: We used the following statistics to evaluate the forecasting accuracy of the alternative models:

1. Mean Error

$$ME_i = \frac{1}{43}\sum_{t=10}^{52} e_{it} \tag{12}$$

2. Mean Absolute Error

$$MAE_i = \frac{1}{43}\sum_{t=10}^{52} |e_{it}| \tag{13}$$

3. Root Mean Squared Error

$$RMSE_i = \sqrt{\frac{1}{43}\sum_{t=10}^{52} e_{it}^2} \tag{14}$$

4. Relative Root Mean Squared Error

$$RRMSE_i = 100 \times \frac{\sqrt{\sum_{t=10}^{52} e_{it}^2}}{\sum_{t=10}^{52} y_{it}} \tag{15}$$

5. Mean Absolute Percentage Error:

$$MAPE_i = \frac{100}{43} \times \sum_{t=10}^{52} \frac{|e_{it}|}{y_{it}} \tag{16}$$

6. Mean Percentage Error:

$$MPE_i = \frac{100}{43} \times \sum_{t=10}^{52} \frac{e_{it}}{y_{it}} \tag{17}$$
