## Supplementary material for "Assessing COVID-19 pandemic excess deaths in Brazil: years 2020 and 2021": S1 Table

| Year | Region | State | COVID-19 | Reported | Expected | Excess | 95% PI | | P-Score | Ratio$_{EC}$ |
|---|---|---|---|---|---|---|---|---|---|---|
| 2020 | N | RO | 1 910 | 9 024 | 7 068 | 1 956 | 1 699 | 2 213 | 27.7 | 1.02 |
| | | AC | 819 | 4 208 | 3 627 | 581 | 367 | 795 | 16.0 | 0.71 |
| | | AM | 6 003 | 21 816 | 15 802 | 6 014 | 5 475 | 6 552 | 38.1 | 1.00 |
| | | RR | 809 | 3 135 | 2 565 | 570 | 385 | 755 | 22.2 | 0.70 |
| | | PA | 7 941 | 45 173 | 35 542 | 9 631 | 8 589 | 10 673 | 27.1 | 1.21 |
| | | AP | 1 117 | 4 031 | 3 120 | 911 | 783 | 1 040 | 29.2 | 0.82 |
| | | TO | 1 181 | 8 022 | 6 984 | 1 038 | 696 | 1 380 | 14.9 | 0.88 |
| | NE | MA | 5 153 | 37 723 | 29 797 | 7 926 | 6 140 | 9 713 | 26.6 | 1.54 |
| | | PI | 2 822 | 20 494 | 17 510 | 2 984 | 2 287 | 3 682 | 17.0 | 1.06 |
| | | CE | 11 883 | 60 363 | 51 109 | 9 254 | 7 498 | 11 011 | 18.1 | 0.78 |
| | | RN | 3 109 | 21 287 | 18 561 | 2 726 | 1 729 | 3 722 | 14.7 | 0.88 |
| | | PB | 3 554 | 26 871 | 23 268 | 3 603 | 2 487 | 4 719 | 15.5 | 1.01 |
| | | PE | 11 499 | 66 627 | 53 765 | 12 862 | 8 047 | 17 676 | 23.9 | 1.12 |
| | | AL | 3 607 | 20 861 | 17 777 | 3 084 | 2 048 | 4 119 | 17.4 | 0.85 |
| | | SE | 2 360 | 13 626 | 11 296 | 2 330 | 1 872 | 2 789 | 20.6 | 0.99 |
| | | BA | 11 041 | 92 747 | 79 306 | 13 441 | 11 377 | 15 505 | 17.0 | 1.22 |
| | SE | MG | 13 672 | 130 816 | 119 271 | 11 545 | 8 401 | 14 689 | 9.7 | 0.84 |
| | | ES | 4 886 | 25 382 | 20 931 | 4 451 | 3 717 | 5 185 | 21.3 | 0.91 |
| | | RJ | 32 257 | 150 480 | 122 534 | 27 946 | 23 119 | 32 773 | 22.8 | 0.87 |
| | | SP | 47 898 | 303 735 | 262 729 | 41 006 | 33 818 | 48 194 | 15.6 | 0.86 |
| | S | PR | 8 848 | 71 329 | 65 464 | 5 865 | 3 225 | 8 505 | 9.0 | 0.66 |
| | | SC | 5 387 | 40 223 | 36 885 | 3 338 | 2 084 | 4 591 | 9.1 | 0.62 |
| | | RS | 9 406 | 79 933 | 78 864 | 1 069 | -2 283 | 4 421 | 1.4 | 0.11 |
| | CW | MS | 2 289 | 16 606 | 14 477 | 2 129 | 1 455 | 2 803 | 14.7 | 0.93 |
| | | MT | 4 412 | 20 513 | 15 895 | 4 618 | 4 025 | 5 210 | 29.1 | 1.05 |
| | | GO | 7 646 | 41 999 | 34 735 | 7 264 | 5 722 | 8 806 | 20.9 | 0.95 |
| | | DF | 3 111 | 14 296 | 10 835 | 3 461 | 3 020 | 3 901 | 31.9 | 1.11 |
| 2021 | N | RO | 4 854 | 13 887 | 8 360 | 5 527 | 5 218 | 5 835 | 66.1 | 1.14 |
| | | AC | 1 291 | 5 467 | 4 415 | 1 052 | 812 | 1 291 | 23.8 | 0.81 |
| | | AM | 8 577 | 28 832 | 19 015 | 9 817 | 9 208 | 10 426 | 51.6 | 1.14 |
| | | RR | 1 218 | 4 239 | 3 259 | 980 | 695 | 1 266 | 30.1 | 0.80 |
| | | PA | 10 356 | 52 017 | 42 538 | 9 479 | 8 169 | 10 789 | 22.3 | 0.92 |
| | | AP | 1 062 | 4 747 | 3 810 | 937 | 793 | 1 081 | 24.6 | 0.88 |
| | | TO | 2 671 | 11 477 | 8 350 | 3 127 | 2 648 | 3 607 | 37.5 | 1.17 |
| | NE | MA | 7 446 | 44 449 | 35 385 | 9 064 | 7 065 | 11 063 | 25.6 | 1.22 |
| | | PI | 4 444 | 25 886 | 20 703 | 5 183 | 4 384 | 5 982 | 25.0 | 1.17 |
| | | CE | 15 665 | 73 459 | 59 239 | 14 220 | 10 294 | 18 145 | 24.0 | 0.91 |
| | | RN | 4 950 | 26 562 | 21 909 | 4 653 | 3 134 | 6 172 | 21.2 | 0.94 |
| | | PB | 5 930 | 34 558 | 27 293 | 7 265 | 5 492 | 9 039 | 26.6 | 1.23 |
| | | PE | 12 716 | 80 406 | 63 153 | 17 253 | 11 837 | 22 669 | 27.3 | 1.36 |
| | | AL | 4 174 | 24 918 | 20 455 | 4 463 | 2 702 | 6 225 | 21.8 | 1.07 |
| | | SE | 3 458 | 16 624 | 13 228 | 3 396 | 2 797 | 3 995 | 25.7 | 0.98 |
| | | BA | 16 867 | 114 698 | 94 655 | 20 043 | 16 955 | 23 131 | 21.2 | 1.19 |
| | SE | MG | 44 198 | 188 518 | 141 739 | 46 779 | 41 854 | 51 704 | 33.0 | 1.06 |
| | | ES | 6 239 | 32 664 | 24 944 | 7 720 | 6 554 | 8 886 | 31.0 | 1.24 |
| | | RJ | 40 531 | 188 007 | 145 351 | 42 656 | 35 780 | 49 531 | 29.4 | 1.05 |
| | | SP | 110 138 | 429 427 | 311 564 | 117 863 | 109 869 | 125 857 | 37.8 | 1.07 |
| | S | PR | 32 824 | 112 182 | 77 921 | 34 261 | 29 653 | 38 869 | 44.0 | 1.04 |
| | | SC | 14 565 | 59 368 | 44 678 | 146 90 | 12 751 | 16 630 | 32.9 | 1.01 |
| | | RS | 26 715 | 116 877 | 95 349 | 21 528 | 15 857 | 27 200 | 22.6 | 0.81 |
| | CW | MS | 6 790 | 24 814 | 17 312 | 7 502 | 6 413 | 8 591 | 43.3 | 1.10 |
| | | MT | 9 172 | 28 476 | 19 014 | 9 462 | 8 802 | 10 122 | 49.8 | 1.03 |
| | | GO | 17 972 | 60 394 | 41 430 | 18 964 | 16 886 | 21 042 | 45.8 | 1.06 |
| | | DF | 5 370 | 18 784 | 12 924 | 5 860 | 5 203 | 6 517 | 45.3 | 1.09 |
